# Phospholipase D3 contributes to Alzheimer’s disease risk via disruption of Aβ clearance and microglia response to amyloid plaques

**DOI:** 10.1101/2022.01.31.22270175

**Authors:** Matthew J. Rosene, Simon Hsu, Shih Feng You, Logan Brase, Anthony Verbeck, Rita Martinez, Clare E. Wallace, Zeran Li, Ping Yan, Nina M. Dräger, Sydney M. Sattler, Abhirami K Iyer, Shannon L Macauley, David M. Holtzman, Bruno A. Benitez, Martin Kampmann, Carlos Cruchaga, Oscar Harari, John R. Cirrito, Jin-Moo Lee, Alison M. Goate, Celeste M. Karch

**Author notes:** Corresponding Author: Celeste M. Karch, Department of Psychiatry, Washington University School of Medicine, 425 S. Euclid Ave, Campus Box 8134, St. Louis, MO 63110.

## Abstract

Alzheimer’s disease (AD) is characterized by the accumulation of amyloid-β (Aβ) plaques and neurofibrillary tangles in the brain. AD is also the result of complex genetic architecture that can be leveraged to understand pathways central to disease processes. We have previously identified coding variants in the *phospholipase D3* (*PLD3*) gene that double the late-onset AD risk. However, the mechanism by which PLD3 impacts AD risk is unknown. One AD risk variant, *PLD3* p.A442A, disrupts a splicing enhancer-binding site and reduces *PLD3* splicing in human brains. Using differentiated induced pluripotent stem cells from a *PLD3* p.A442A carrier and CRISPR-reverted, isogenic control, we show that *PLD3* p.A442A cortical neurons exhibit a *PLD3* splicing defect and a significant increase in Aβ42 and Aβ40, both of which are corrected upon reversion of the risk allele in isogenic control neurons. Thus, *PLD3* p.A442A is sufficient to alter *PLD3* splicing and Aβ metabolism. While the normal function of PLD3 is poorly understood, *PLD3* is highly expressed in neurons and brain regions most susceptible to amyloid pathology. *PLD3* expression is significantly lower in AD brains than controls, suggesting that PLD3 may play a role in sporadic AD. Thus, we sought to determine whether PLD3 contributes to Aβ accumulation in AD. In a mouse model of amyloid accumulation, loss of *Pld3* increases interstitial fluid (ISF) Aβ and reduces Aβ turnover. AAV-mediated overexpression of *PLD3* in the hippocampus decreased ISF Aβ levels and accelerated Aβ turnover. To determine whether PLD3-mediated reduction of ISF Aβ impacts amyloid accumulation, we measured amyloid plaque abundance and size after significant Aβ deposition. We found that in the absence of *Pld3*, amyloid plaques were less compact and more diffuse. Additionally, we observed reduced recruitment of microglia to amyloid plaques in the absence of *Pld3*. PLD3 may impact amyloid accumulation and AD risk through disrupted microglia function as *PLD3* is enriched in disease associated microglia in human brains. Together, our findings demonstrate that PLD3 regulates Aβ clearance through cell-autonomous and non-cell-autonomous pathways in a manner that likely contributes to AD risk.

## Introduction

Alzheimer’s disease (AD) is pathologically defined by neuronal loss and the accumulation of amyloid-β (Aβ) plaques and neurofibrillary tangles in the brain. Genetic, biochemical, and neuropathologic data suggest that Aβ aggregation is central to initiating AD pathogenesis [1]. Rare mutations in *APP*, *PSEN1,* and *PSEN2* cause dominantly inherited AD. Late-onset AD (LOAD) also has a strong genetic component [2]. Identifying novel loci that affect LOAD risk is critical to our understanding of the underlying etiology of AD and novel therapeutic pathways.

The genetic architecture that underlies LOAD is complex [2]. Large-scale genomic studies have led to the identification of novel genes and risk loci that contribute to AD [2, 3]. Whole exome sequencing of densely affected LOAD families has revealed a rare genetic variant within *PLD3* (p.V232M) that perfectly segregated with disease in two independent families and doubled AD risk in seven independent case-control series (4,998 AD cases/6,356 controls; OR=2.10, p=2.93×10^-5^) [4]. Gene-based analyses indicated that multiple variants in *PLD3* increase AD risk in European (EA) and African American (AA) populations (e.g.: p.M6R, p.V232M, p.A442A; EA: OR=2.75, p=1.44×10^-11^; AA: OR=5.48, p=1.40×10^-3^) [4].

PLD3 is a non-classical member of the phospholipase D (PLD) family of enzymes, with no known function [5–7]. *PLD3* is expressed in pyramidal neurons within the brain, and in AD brains, PLD3 co-localizes with amyloid plaques [8, 9]. Common variants in *PLD3* are associated with CSF Aβ levels, an AD biomarker [10]. *In vitro*, *PLD3* expression is correlated with extracellular Aβ levels: *Pld3* silencing is associated with increased Aβ levels, and *PLD3* overexpression is associated with reduced Aβ levels [4]. Thus, PLD3 may play a broader role in LOAD.

Here, we sought to define the contribution of *PLD3* risk variants to AD-related phenotypes in human stem cell models and the role of PLD3 in amyloid pathology in mouse models. We found that a *PLD3* risk variant is sufficient to increase Aβ levels in stem cell-derived neurons. In animal models of amyloid accumulation, *Pld3* silencing reduces Aβ turnover and alters the composition of amyloid plaques. In the absence of *Pld3*, microglia recruitment to plaques is attenuated. Together, this study suggests that PLD3 contributes AD pathogenesis via Aβ clearance through cell-autonomous and non-cell-autonomous pathways.

## Materials and Methods

### Patient Consent

Skin biopsies were collected following written informed consent from the donor. The Washington University School of Medicine Institutional Review Board and Ethics Committee approved the informed consent (IRB 201104178, 201306108). The consent allows for the use of tissue by all parties, commercial and academic, for research but not for human therapy.

### Dermal Fibroblast Isolation

Dermal fibroblasts were isolated from skin biopsies obtained from the Knight Alzheimer Disease Research Center (ADRC) research participants. Briefly, skin biopsies were collected by surgical punch and stored in Fibroblast Growth Media (Lonza). To isolate dermal fibroblasts from a skin biopsy, the biopsies were rinsed with PBS and cut lengthwise with dissecting scissors. The resulting tissue sections were then plated into a dry 24-well tissue culture-treated plate (approximately 6-12 sections). After removing excess PBS from the wells, 300ul of fibroblast growth media (Lonza) was carefully added and tissue was incubated at 37°C and 5% CO_2_. After 24 hours, tissue was supplemented with 1mL fibroblast growth media and media changes were repeated every 3-4 days. Fibroblast cells migrated from the tissue within two weeks of culture. Dermal fibroblasts were maintained in fibroblast growth media (Lonza) supplemented with penicillin/streptomycin.

### iPSC Generation, Characterization, and Maintenance

Human fibroblasts (F13504) were transduced with non-integrating Sendai virus carrying the four factors required for reprogramming: OCT3/4, SOX2, KLF4, and cMYC [11, 12]. Cells showing morphological evidence of reprogramming were selected by manual dissection.

Human iPSCs were cultured using feeder-free conditions (Matrigel, BD Biosciences, Franklin Lakes, NJ, USA). Human iPSCs were thawed (1-2 x 10^6^ cells/mL), diluted in DMEM/F12, and centrifuged at 750 rpm for 3 minutes. The resulting iPSC pellet was then diluted in mTeSR1 supplemented with Rock inhibitor (Y-27632; 10µM final). IPSC were subsequently cultured in 37°C, 5% CO_2_ with daily medium changes (mTesR1, STEMCELL Technologies, Vancouver, BC, CA).

All iPSC lines were characterized using standard methods [11]. Each line was analyzed for pluripotency markers (OCT4A, SOX2, SSEA4, TRA1-80) by immunocytochemistry (ICC) (Invitrogen A24881) and quantitative PCR (qPCR); for chromosomal abnormalities by karyotyping; and for *PLD3* variant status by Sanger sequencing (Supplemental Figure 1).

### Genome editing

We used CRISPR/Cas9 to generate isogenic control lines for the *PLD3* p.A442A iPSC as previously described [13]. The p3s-Cas9HC Cas9 expression plasmid (Addgene 43945) and CRISPR reagents (Addgene plasmid 43860) were used [14]. Guide RNAs were designed to overlap with the allele to be modified and have at least 3bp of mismatch to any other gene in the human genome. The activity of the guide was validated using a mismatch detection assay to determine non-homologous end-joining efficiency in K562 cells. A correctly edited clone and an unmodified clone were identified, expanded and characterized as described above for karyotyping and pluripotency markers. Sanger sequencing was performed for the on-target and predicted off-target sites.

### Cortical neuron differentiation

IPSCs were differentiated into neuronal cells using a two-step approach as previously described [13]. IPSCs were plated at a density of 65,000 cells per well in neural induction media (STEMCELL Technologies) in a 96-well v-bottom plate to form highly uniform neural aggregates and, after five days, transferred onto culture plates. The resulting neural rosettes were then isolated by enzymatic selection (Neural Rosette Selection Reagent; STEMCELL Technologies) and cultured as neural progenitor cells (NPCs). NPCs were differentiated in planar culture in neuronal maturation medium (neurobasal medium supplemented with B27, GDNF, BDNF, cAMP). Neurons typically arise within one week after plating, identified using immunocytochemistry for β-tubulin III (Tuj1). The cells continued to mature and were analyzed at six weeks.

### Immunocytochemistry

IPSC-derived neurons were grown on PLO/laminin-coated 8-well chamber slides (Millipore). Culture media was aspirated, and cells were then washed and fixed with 4% paraformaldehyde (Sigma). After several washes, cells were permeabilized with 0.1% Triton X-100 in PBS. Cells were then blocked in 0.1% bovine serum albumin (BSA; Sigma) and treated with primary (Tuj1) and secondary antibodies diluted in 0.1% BSA. Immunostained cells were then imaged (Nikon Eclipse 80i fluorescent microscope).

### Transcriptomics and Digital Deconvolution

RNA was extracted from iPSC-derived neurons from *PLD3* p.A442A vs. the isogenic controls using Tissue Lyser LT and RNeasy Mini Kit (Qiagen, Hilden, Germany). RNA-seq paired-end reads with read lengths of 2 × 150 bp were generated using Illumina HiSeq 4000 with a mean coverage of 80 million reads per sample. FastQC was applied to perform quality control and aligned to human GRCh37 primary assembly using Star (ver 2.5.2b). We applied Salmon transcript expression quantification (ver 0.7.2) to infer the gene expression. To estimate the relative proportion of major brain cell types in the dish, digital deconvolution was performed as previously described using a reference marker panel [15].

### PLD3 Splicing Assay

RNA was extracted from cell lysates with an RNeasy kit (Qiagen) according to the manufacturer’s protocol. Extracted RNA (10ug) was converted to cDNA by PCR using the High-Capacity cDNA Reverse Transcriptase kit (ABI). SYBR-green primers were designed using Primer Express software, Version 3 (ABI). Real-time PCR assays were used to quantify *PLD3* exon 7 (forward primer: GCAGCTCCATCCCATCAACT; reverse: CTTGGTTGTAGCGGGTGTCA), exon 8 (forward primer: CTCAACGTGGTGGACAATGC; reverse: AGTGGGCAGGTAGTTCATGACA), exon 9 (forward primer: ACGAGCGTGGCGTCAAG; reverse: CATGGATGGCTCCGAGTGT), exon 10 (forward primer: GGTCCCCGCGGATGA; reverse: GGTTGACACGGGCATATGG) and exon 11 (forward primer: GCTGCTGGTGACGCAGAAT; reverse: AGTCCCAGTCCCTCAGGAAAA). Each qPCR analysis included technical replicates and biological triplicates. Real-time data were analyzed using the comparative Ct method. Only samples with a standard error of <15% were analyzed. The Ct values for exon 11 were normalized with the Ct value for exons 7-10. The relative exon 11 levels for the iPSC-derived neurons from *PLD3* p.A442A vs. the isogenic control were compared using a Tukey’s t-test.

### Aβ Measurements

Conditioned medium was collected from neurons after six weeks in culture and centrifuged at 3,000xg at 4°C for 10 minutes to remove cell debris. The levels of Aβ42 and Aβ40 were measured in cell culture media by sandwich ELISA as described by the manufacturer (Life Technologies). To account for variability in transfection efficiency between experiments, ELISA values were obtained (pg/mL) and corrected for total intracellular protein (ug/mL). Statistical difference was measured using an unpaired Student’s t-test.

#### PLD3 expression in AD and neuropathology free human brains

Laser microdissected neurons from AD and control brains were previously reported and deposited as GSE5281 [38]. Brain samples were analyzed from 47 individuals of European descent clinically and neuropathologically confirmed AD cases or controls. The 33 AD samples were 54.5% female with a mean age of 79.9 years (range 73–86.8) and an average postmortem interval (PMI) of 2.5 hours. The 14 control brains were 28.6% female with a mean age of 79.8 years (range 70.1–88.9). Samples were obtained from the entorhinal cortex, hippocampus, medial temporal gyrus, posterior cingulate, superior frontal gyrus, and primary visual cortex. RNA expression was measured using an Affymetrix GeneChip for gene expression. The log-transformed expression values were analyzed with brain region, age, and sex as covariates to analyze RNA expression.

Gene expression was analyzed in a second, independent, publicly available dataset of the temporal cortex of 76 control and 80 AD brains (syn6090813). Differential gene expression comparing controls v. AD was performed using a “Simple Model”, multi-variable linear regression analyses were conducted in R, using normalized gene expression measures including sex, age-at-death, RNA integrity number, brain tissue source, and flowcell as covariates [16]. Statistical difference was measured using an unpaired Student’s t-test.

### Single nuclei RNAseq in human brains

Human parietal cortices were processed to isolate nuclei, and the nuclei were then sequenced using the 10X Chromium single cell Reagent Kit v3, with 10,000 cells per sample and 50,000 reads per cell for each of the 74 samples as previously described [17, 18]. The CellRanger (v3.0.2 10XGenomics) software was used to align the sequences and quantify gene expression. We used the GRCh38 (3.0.0) reference to prepare a pre-mRNA reference. Filtering and QC were done using the Seurat package (3.0.1) on each subject individually. After the Uniform Manifold Approximation and Projection (UMAP) analysis was performed with the top 14 PCs, we then used Seurat *FindNeighbors* and *FindClusters* functions to identify unique cell states or subclusters. Samples included in the subsequent analyses are summarized in Supplemental Table 1. To identify associations between cell-type transcriptional state and disease status or genetic strata (control, sporadic AD (sAD), TREM2), we applied linear regression models to test the cell state compositions of each subject. The proportions were normalized using a cube root transformation and were corrected by sex and age of death.

Differentially expressed genes among the individual cell states were identified using a linear mixed model that corrected for sex and subject. To determine if there was unique functionality or potentially altered expression levels associated with disease/genetic carriers in the alternative cell states, we employed linear mixed model that predicted the expression level of each gene, modeled as zero-inflated negative binomial distributions and corrected for sex and age of death. Donors were modeled as random variables as previously described [17]. Data can be publicly accessed at http://ngi.pub/SNARE.

### Mouse Models

Animal care and surgical procedures were approved by the Animal Studies Committee of Washington University School of Medicine in accordance with guidelines of the United States National Institutes of Health. APPswe/PS1ΔE9 transgenic mice (APP/PS1; The Jackson Laboratory; 034829) [19] of both sexes were used in this study. APP/PS1 mice were maintained on a C57bl6;C3B6 mixed background.

*Pld3-KO* mice were generated using CRISPR/Cas9 technology. gRNAs were designed to target an early conserved exon (mPld3.g19; TGCTGTGAGCACCGGCAAGGNGG). Guide activity was assessed in mouse neuroblastoma cells (N2A) using a T7E1 mismatch detection assay as previously described [13]. RNA was injected into the pronuclei of fertilized, viable murine oocytes isolated from a set of C57Bl/6xCBA (hybrid) female mice. Founders were identified using mPLD3 screening primers SM406.Cel.F 5’ CATGGGCACTGTATCCCATCT 3’ and SM406.Cel.R 5’ AGGACACAAAAACGTCACCCT 3’, which generated a parental band (575bp) and fragments (307 and 268bp). Subsequent generations were backcrossed to C57Bl/6;C3B6 (Supplemental Figure 2).

APP/PS1 mice were crossed with the *Pld3*-KO mice to generate APP/PS1x*Pld3*^+/-^. APP/PS1x*Pld3*^+/-^ were crossed to obtain APP/PS1x*Pld3*-KO (APP/PS1x*Pld3*^-/-^) mice and APP/PS1x*Pld3*-WT (APP/PS1x*Pld3*^+/+^) littermates. Animals of both sexes were used in the study.

### Intracranial AAV-mediated expression of PLD3 and shPLD3

Adeno-associated virus (AAV8) particles were generated that express AAV-CMV-hPLD3-WT-GFP, AAV-CMV-GFP (control), AAV-U6-shPLD3-GFP, and AAV-U6-shScrambled-GFP (control). Viral particles were injected into the hippocampus of APP/PS1 mice of both sexes to produce widespread transduction in hippocampal neurons as previously described [20, 21]. Two uL of AAV particles (1.5 × 10^12^ vg/ml) were stereotaxically injected bilateral hippocampi (2μl over 10 minutes) in 3-month-old APP/PS1 transgenic mice (prior to plaque appearance) [22, 23]. Virus is detectable at least 4-5 months post-injection [21, 24].

### In Vivo Microdialysis

Prior to plaque accumulation (5 months of age) and two months post AAV8-injection, hippocampal ISF Aβ levels were quantified using microdialysis in APP/PS1 mice as previously described [25, 26]. The use of a 38-kDa MWCO semi-permeable membrane allows for molecules smaller than this cut-off to diffuse into the probe. The probe was flushed with perfusion buffer at a constant rate (1.0μl/minute) and collected into a refrigerated fraction collector, and then assayed by sandwich ELISA (Aβ_x-40_). A mouse monoclonal anti-Aβ_40_ capture antibody (mHJ2) made in-house was used in conjunction with a biotinylated central domain detection antibody (mHJ5.1) and streptavidin-poly-HRP-40 (Fitzgerald Industries, Acton, MA)[27]. During microdialysis, mice were housed in cages that permit free movement and ad-libitum food/water, while ISF Aβ was sampled. Baseline levels of ISF Aβ were sampled every 60 minutes between hours 5-12 (after the microdialysis probe insertion) and averaged to determine the baseline ISF Aβ levels in each mouse. At hour 12, mice were administered the γ-secretase inhibitor Compound E (20mg/kg), intraperitoneally, which enabled us to determine the elimination half-life of ISF Aβ [28]. Aβ half-life was calculated using first-order kinetics. Microdialysis was performed with sham and littermate controls. Statistical difference was measured using an unpaired Student’s t-test.

To evaluate the impact of *Pld3* on Aβ kinetics, APP/PS1x*Pld3*-WT and APP/PS1x*Pld3*-KO mice were implanted at one-month post-injection with a microdialysis probe with a 38-kDa MWCO probe, and ISF was collected as described above. After 12 hours, γ-secretase inhibitor LY411575 (3mg/kg in 50:50PBS:PEG400) was administered intraperitoneally to define the elimination half-life of ISF Aβ. Aβ half-life was calculated as described above. Microdialysis was performed with sham and littermate controls. Statistical difference was measured using an unpaired Student’s t-test.

### Brain Tissue Preparation

APP/PS1x*Pld3*-WT and APP/PS1x*Pld3*-KO mice were anesthetized with sodium pentobarbital and perfused with 0.3% heparin in PBS at nine months of age. Brains were dissected and cut into two hemispheres. The right hemisphere was snap-frozen on dry ice and stored at 80°C for biochemical analyses. The left hemisphere was fixed in 4% paraformaldehyde for 24 hours, followed by 30% sucrose in PBS at 4°C. Coronal sections (50 µm) were cut on a freezing-sliding microtome. Collected slices were stored in cryoprotectant solution (0.2 M phosphate-buffered saline, 30% sucrose, and 30% ethylene glycol) at −20°C.

### Immunohistochemistry

APP/PS1x*Pld3*-WT and APP/PS1x*Pld3*-KO brains were cut into 50µm sections from the rostral anterior commissure to the caudal hippocampus. Brains sections (n=20 mice per group) were incubated in 0.3% H_2_O_2_ in Tris-buffered saline (TBS) for 10 minutes and blocked in 3% milk in TBS-X for 30 minutes. Tissue was incubated in HJ3.4B antibody (anti-Aβ-1-13; 2.4 µg/ml; a generous gift from the Holtzman lab) overnight [29]. Sections are then incubated in Vectastain ABC elite solution in TBS (1:400) for 1 hour before incubating in 3,3’-Diaminobenzidine (DAB) solution. Sections were then mounted onto slides with Cytoseal (Thermo Scientific 8312-4). A Nanozoomer Digital Scanner (Hamamatsu Photonics) was used to create high-resolution digital images for the HJ3.4-stained brain slices. Total plaque coverage, total plaque count, and average plaque size were analyzed using NIH ImageJ software. Total plaque coverage was expressed as a percentage of the total area for each slice and averaged across the three slices per animal. Plaque count was expressed as the plaque count per nm^2^ averaged across the three slices per animal. Plaque size was expressed in µm^2^ averaged across all three slices for each mouse. Both the hippocampus and cortex were analyzed separately for each slice. Statistical difference for plaque size, percentage of total area, and plaque count per square millimeter was measured using an unpaired Student’s t-test.

### X-34 Plaque Staining

To evaluate X-34 positive plaque burden, frozen coronal brain sections (50 µm) were mounted on Superfrost Plus slides, permeabilized with 0.25% Triton X-100 in PBS for 30 minutes and stained with X-34 (0.01mM; Sigma SML1954) dissolved in 40% ethanol in PBS, pH 10 for 20 min. The tissue was then washed with 40% ethanol in PBS and mounted with Fluoromount G mounting medium. Slides were imaged using Cytation7 (BioTek) and quantified using NIH ImageJ software. A Cytation7 cell imaging multi-modal imager was used to create high-resolution images of the X-34 scanned brain sections. Total plaque coverage, total plaque count, and average plaque size were analyzed using NIH ImageJ software. Total plaque coverage was expressed as a percentage of the total area for each section and averaged across three sections per animal. Plaque count was expressed as the plaque count per square millimeter averaged across the three sections per animal. Plaque size was expressed in square microns averaged across all three sections for each mouse. Both the hippocampus and cortex were analyzed separately for each section. Statistical difference for plaque size, percentage of total area, and plaque count per square millimeter was measured using an unpaired Student’s t-test.

### Immunofluorescence

Coronal brain sections (50µm) were permeabilized in 0.25% Triton X-100 for 30 min at room temperature. Slices were then stained with X-34 (0.001 mM; Sigma SML1954) dissolved in 40% ethanol in PBS for 20 min and blocked in 3% BSA/3% Normal Donkey serum in 0.1% Triton X-100 in PBS. X-34-stained samples were immunostained with antibodies HJ3.4 (2.4ug/mL; α-Aβ1-13) or Iba1 (Abcam ab5076) and CD68 (BioRad MCA1957) at 4°C overnight. Three slices per animal were stained. Secondary antibodies of Alexa Fluor 488-conjugated donkey anti-rat IgG, Alexa Fluor 647-conjugated donkey anti-goat, and Alexa Fluor 647 donkey anti-mouse (Invitrogen). Slides were mounted with Fluoromount G mounting media. High resolution 40x images were obtained using the Zeiss LSM 880 Confocal microscope with Airyscan. Z-stacks were analyzed using NIH ImageJ software, where max projection images (those with the highest signal intensity) were selected for each fluorescent channel. The Iba1 and CD68 signals were quantified within 15µm of the X-34-positive plaques. Iba1 and CD68 colocalization was also quantified. An outlier analysis was run using the ROUT method with a Q-value of 1%. Three outliers were removed from the APP/PS1x*Pld3*-KO mice in the Iba1 analysis. Statistical difference was measured using an unpaired Student’s t-test.

To quantify HJ3.4 and X-34 colocalization, Z-stacks were analyzed using NIH ImageJ software, where max projection images were selected for each fluorescent channel. Plaque composition was quantified by measuring the percentage of HJ3.4 staining within X-34-positive areas. Nonfibrillar plaque area was normalized to the total X-34 area. A ROUT outlier analysis with a Q-value of 1% was run for each of these quantifications. No outliers were removed from the analysis of percent area. Four outliers were excluded in the APP/PS1x*Pld3*-WT mice, and three outliers were excluded in the APP/PS1x*Pld3*-KO mice in the nonfibrillar plaque area analysis. Statistical difference was measured using an unpaired Student’s t-test.

## Results

### AD risk variant PLD3 p.A442A is sufficient to alter PLD3 splicing and Aβ levels

Three highly conserved, rare variants in *PLD3* increase AD risk: p.M6R, p.V232M, and p.A442A [4]. *PLD3* p.A442A (G>A) was associated with increased AD risk in four independent case-control datasets (p=3.78×10-7; OR=2.12) [4]. *PLD3* p.A442A was predicted to modify a splicing enhancer-binding site, SRSF1, *in silico* [4]. In the brains of *PLD3* p.A442A carriers, total *PLD3* and exon 11-containing transcript expression were reduced compared to controls [4]. However, these association studies did not allow us to attribute the causality of the risk variant to the phenotype.

Here, we coupled genome-editing with stem cell models to determine whether *PLD3* p.A442A is sufficient to alter *PLD3* splicing and phenocopy AD-related phenotypes. Primary dermal fibroblasts were obtained from a patient carrying a single copy of the *PLD3* p.A442A variant. Fibroblasts were de-differentiated into induced pluripotent stem cells using non-integrating Sendai virus. The resulting iPSC were characterized for pluripotency markers, the presence of the *PLD3* p.A442A variant, and chromosomal integrity (Supplemental Figure 1). To determine the causality of *PLD3* p.A442A on AD-related phenotypes, we used CRISPR/Cas9 to correct the A allele to G (wild-type) (Figure 1A) [13]. *PLD3* p.A442A and isogenic controls (*PLD3* WT) were then differentiated into cortical neurons using a growth factor-mediated approach as previously described (Figure 1A) [13], where they illustrated a similar capacity to form Tuj1-positive neurons (Figure 1B). To further verify the similarity between the cells in their capacity to form neurons, we estimated the relative proportion of neurons in the cultures using bulk transcriptomics and deconvolution methods that include all the major brain cell types [15]. We found that differentiated cultures from *PLD3* p.A442A and isogenic controls exhibited a similar enrichment of neurons (∼95%; Figure 1C). These findings are consistent with a variant that impacts a late-onset disease, where we would not predict a significant developmental defect.

**Figure 1:**
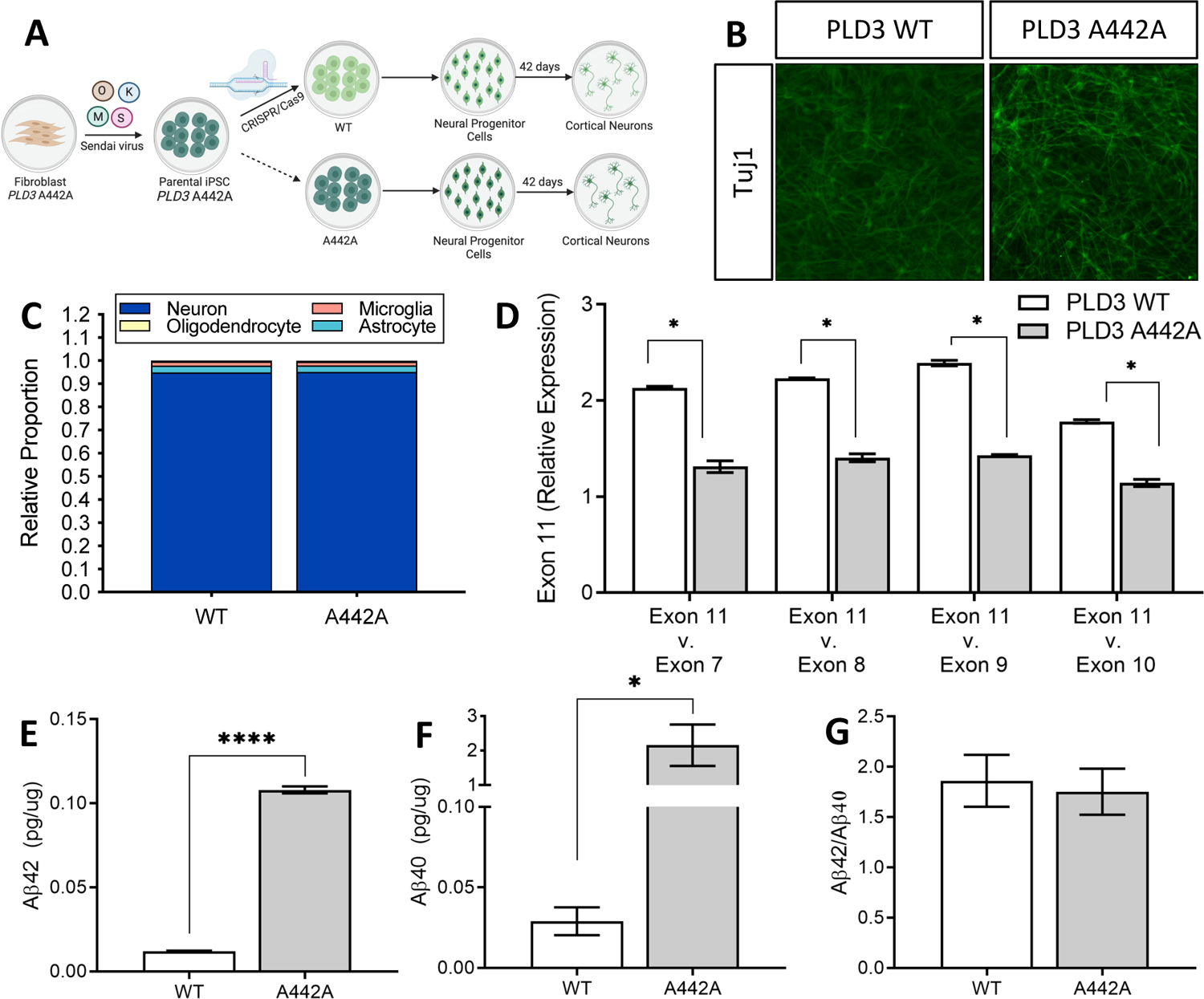
iPSC-neurons expressing *PLD3* p.A442A phenocopy splicing defects observed in human brains. A. Fibroblasts from a *PLD3* p.A442A variant carrier were reprogrammed into induced pluripotent stem cells (iPSCs). CRISPR-Cas9 technology was used to generate an isogenic control line. IPSCs were then differentiated into cortical neurons (see Methods). Downstream assays were performed after 42 days in culture. B. iPSC-derived neurons stained with Tuj1 illustrate a similar capacity of p.A442A and isogenic controls (*PLD3* WT) to form neurons. C. Digital deconvolution of iPSC-derived neurons from transcriptomic data illustrates a similar enrichment of neurons in *PLD3* p.A442A and isogenic controls *(PLD3* WT). D. Expression of *PLD3* exon 11 compared to *PLD3* exons 7, 8, 9, and 10. E-G. Sandwich ELISA of media from iPSC-derived neurons (pg/mL) and corrected for total protein measured by BCA (pg/µg). Aβ42 (E), Aβ40 (F), Aβ42/40 (G). Graphs represent mean ± SEM. *<0.05, ****<0.00005. Analyzed by two-tailed Student’s *t* test.

Having demonstrated that *PLD3* p.A442A and isogenic controls share a similar capacity to form neurons, we next asked whether the *PLD3* p.A442A variant was sufficient to alter *PLD3* splicing in a manner consistent with our prior observations in human brains from *PLD3* p.A442A carriers [4]. RNA isolated from neurons expressing *PLD3* p.A442A and isogenic controls was converted to cDNA, and *PLD3* exons 7, 8, 9, 10, and 11 were amplified and quantified. We found that *PLD3* exons are significantly reduced in the *PLD3* p.A442A neurons, which is restored upon correction of the variant allele to WT (Figure 1D). To determine whether *PLD3* p.A442A neurons exhibit changes in Aβ, which would be consistent with a variant that impacts amyloid plaque deposition, we measured extracellular Aβ by sandwich ELISA in the media of *PLD3* p.A442A and isogenic control neurons. After correcting for total protein, we observed a significant increase in Aβ42 and Aβ40 in media from neurons expressing *PLD3* p.A442A (Figure 1E and 1F) without changing the Aβ42/40 ratio (Figure 1G) when compared to isogenic controls. Together, these findings illustrate that in nearly identical, isogenic neurons, the presence of the *PLD3* p.A442A is sufficient to alter *PLD3* transcripts and extracellular Aβ levels.

### PLD3 expression is altered in LOAD brains

Having demonstrated that the AD risk variant in *PLD3* (*PLD3* p.A442A) was sufficient to alter *PLD3* transcripts and Aβ levels, we sought to determine whether *PLD3* is altered in LOAD. We examined *PLD3* expression in laser-captured microdissected neurons across multiple brain regions from AD cases and neuropathology-free controls. *PLD3* expression was significantly lower in AD brains compared with control brains in the entorhinal cortex, hippocampus, medial temporal gyrus, and superior frontal gyrus (Figure 2), regions that exhibit amyloid and tau pathology. Interestingly, *PLD3* expression was unaltered in the primary visual cortex, which is largely spared of AD pathology (Figure 2) [30]. *PLD3* expression was also significantly reduced in an independent cohort of temporal cortices isolated from AD and control brains (β=-0.36; p=3.23×10^-4^) [16]. These findings are consistent with prior reports of reduced *PLD3* expression in LOAD brains [4].

**Figure 2:**
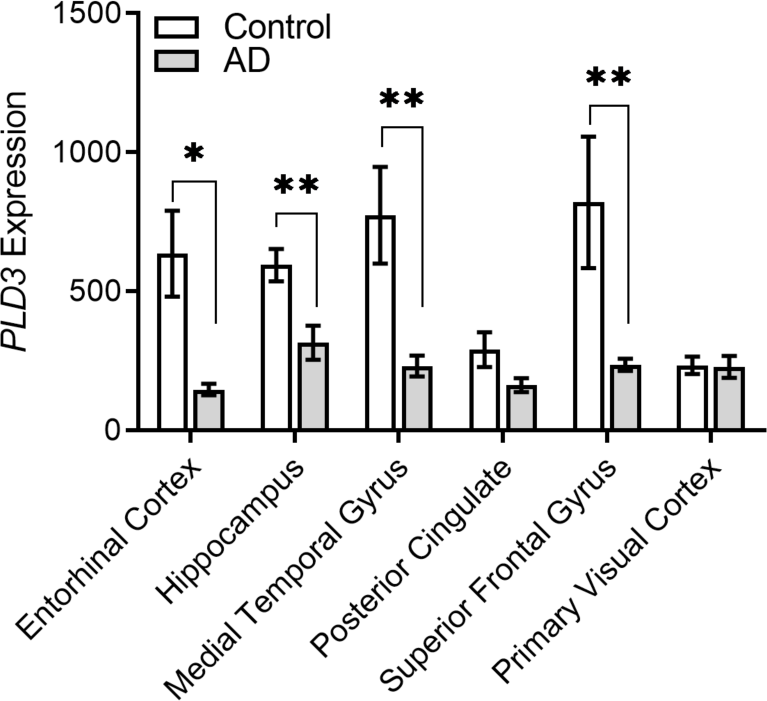
*PLD3* expression is significantly reduced in brain regions vulnerable to AD pathology. Laser capture of microdissected neurons from the brains of neuropathology confirmed control and AD brains [54]. Quantification of *PLD3* expression in laser microdissected neurons isolated from AD and control brains. The graph represents mean ± SEM. *p<0.05. Analyzed by two-tailed Student’s *t* test.

### Pld3 regulates Aβ in APP/PS1 mice

*PLD3* expression is reduced in brains from *PLD3* p.A442A carriers and in LOAD brains (Figure 2) [4], and overexpression or silencing of PLD3 in mouse neuroblastoma cells leads to inverse changes in Aβ levels [4]. Thus, we sought to determine whether modulating *PLD3* expression is sufficient to alter Aβ *in vivo*. In mice, Aβ is primarily generated in neurons and released into the ISF, where it can be cleared by extracellular proteolysis, transported into CSF or across the blood-brain barrier, or by cellular uptake and degradation. The steady-state level of ISF Aβ, thus, reflects these production and degradation/clearance mechanisms. We hypothesized that reducing endogenous *Pld3* expression, as observed in human AD brains, would elevate ISF Aβ levels. To address this hypothesis, 3-month-old APP/PS1 mice were injected with AAV8 particles containing shPld3 or shScrambled (control) and evaluated at five months of age by *in vivo* microdialysis (Figure 3A). At five months of age, shPld3 was sufficient to significantly reduce endogenous *Pld3* transcript level in the hippocampus compared with scrambled controls by 28% (Figure 3B). This modest reduction of *Pld3* did not alter steady-state ISF Aβ levels (Figure 3C and 3D; p=0.27). Next, to test the impact of *Pld3* silencing on the Aβ elimination rate (half-life), Aβ levels were monitored after treatment with a γ-secretase inhibitor (Figure 3C). The secretase inhibitor rapidly blocks Aβ generation within minutes, then ISF is sampled hourly to calculate the rate of elimination of existing Aβ. Silencing of *Pld3* resulted in a 135% increase in Aβ elimination half-life (Figure 3C and 3E; p=0.0025). Thus, PLD3 is likely involved in Aβ clearance.

**Figure 3:**
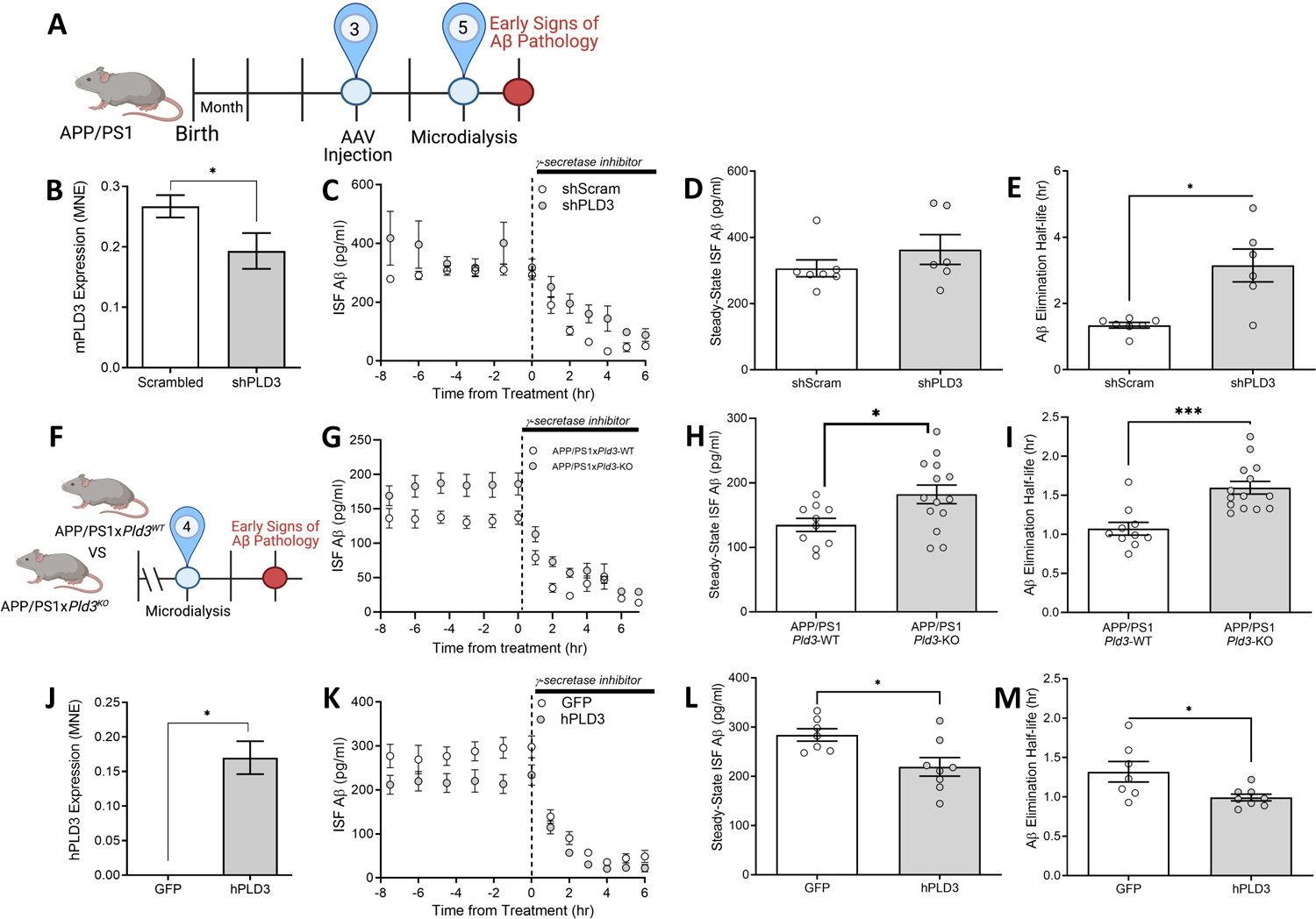
Bi-directional expression of *Pld3* alters Aβ turnover *in vivo*. A-E. The impact of *Pld3* silencing on ISF Aβ. A. Diagram of the experimental timeline: APP/PS1 mice were injected with shScramble shPld3 (compared to *shScram* control)-containing AAV8 particles at three months of age and were evaluated by *in vivo* microdialysis at five months of age. *shScramble* (n=7) and *shPld3* (n=6). B. Knockdown of endogenous *Pld3*. C. Aβ levels in ISF sampled over 14 hours in *shScram,* and *shPld3* injected APP/PS1 mice. D. Steady-state levels of ISF Aβ. E. Elimination half-life of ISF Aβ. F-I. The impact of *Pld3* KO on ISF Aβ. F. Diagram outlining the experimental timeline: APP/PS1x*Pld3*-WT and APP/PS1x*Pld3*-KO mice were evaluated by *in vivo* microdialysis at four months of age. APP/PS1x*Pld3*-WT (n=10), APP/PS1x*Pld3*-KO (n=14). G. Aβ levels in ISF sampled over 14 hours in APP/PS1x*Pld3*-WT and APP/PS1x*Pld3*-KO mice. H. Steady-state levels of ISF Aβ. I. Elimination half-life of ISF Aβ. J-M. APP/PS1 mice were injected with hPLD3 (compared to GFP control)-containing AAV8 particles at three months of age and were evaluated by *in vivo* microdialysis at five months of age. GFP (n=7), hPLD3 (n=8). J. Overexpression of *hPLD3*. K. Aβ levels in ISF sampled over 14 hours in *GFP* and *hPLD3* injected APP/PS1 mice. L. Steady-state levels of ISF Aβ. M. Elimination half-life of ISF Aβ. Graphs represent mean ± SEM. *<0.05, ***<0.0005. Analyzed by two-tailed Student’s *t* test.

The striking impact of a modest *Pld3* decrease in Aβ levels in the APP/PS1 mice led us to investigate the impact of a global knockout of *Pld3* on Aβ (Figure 3F). Global *Pld3* knockout mice were generated using CRISPR/Cas9. A guideRNA targeting an early, highly conserved exon was validated *in vitro* and injected into murine oocytes (see Methods; Supplemental Figure 2). Founders were established and backcrossed to C57Bl/6;C3B6 prior to breeding with APP/PS1 mice (Supplemental Figure 3C). Consistent with prior reports, *Pld3* KO mice were viable and did not exhibit gross defects [9, 31]. *Pld3*-deficient APP/PS1 mice exhibited a 35% increase in steady-state ISF Aβ at four months of age (Figure 3G-3I). In agreement with the AAV-mediated knockdown, *Pld3*-deficient APP/PS1 mice exhibited a 49% increase in Aβ elimination half-life following the administration of a γ-secretase inhibitor (Figure 3G and 3I). Thus, *Pld3* reduction in the brain is sufficient to reduce the turnover of ISF Aβ.

*In vitro* overexpression of *PLD3* was sufficient to reduce extracellular Aβ [4]. Thus, we asked whether overexpression of *hPLD3* in APP/PS1 mice could rescue the ISF Aβ phenotype (Figure 3A). The hippocampus of APP/PS1 mice was bilaterally injected with AAV8 particles containing hPLD3 or GFP (control) at three months of age, and two months later, ISF Aβ was measured by microdialysis (Figure 3A). Overexpression of *hPLD3* significantly reduced steady-state levels of ISF Aβ and Aβ elimination half-life by approximately 25% (Figure 3J-M). Taken together, our findings illustrate that PLD3 expression regulates Aβ turnover in APP/PS1 mice.

### Pld3-deficiency alters plaque composition

Impairment in protein clearance has been implicated in amyloid plaque accumulation and AD pathogenesis [32, 33]. Aβ aggregation in the extracellular space (ISF) into soluble oligomers or insoluble amyloid plaques is a critical driver of AD pathogenesis, and conversion of monomeric Aβ into these aggregates is facilitated at higher concentrations [34]. Thus, we sought to determine whether increased ISF Aβ in four months old APP/PS1x*Pld3*-KO mice could impact amyloid plaque pathology in older animals (Figure 4A). To assess plaque pathology, APP/PS1x*Pld3-*KO mice were sacrificed at nine months of age, and brain sections were co-stained with HJ3.4 (total Aβ) and X-34 (β-sheet rich dense cores; Figure 4B). Plaque composition was then analyzed as the percent of X-34 stain within HJ3.4-positive plaques (termed: fibrillar plaques) and the extent of HJ3.4-positivity outside X-34 plaques (termed: Non-fibrillar plaque area). In APP/PS1x*Pld3*-KO mice, the percentage of fibrillar plaques was significantly reduced compared with APP/PS1xPld3-WT mice (Figure 4C). Conversely, the non-fibrillar plaque area was significantly increased compared with APP/PS1xPld3-WT mice (Figure 4D). In complementary analyses, we found that Aβ plaque size was significantly increased in the cortex of APP/PS1x*Pld3*-KO compared with APP/PS1x*Pld3-*WT mice (Supplemental Figure 3) without a change in the overall plaque burden as defined by the percentage area of HJ3.4-positive immunostaining (e.g., plaque density). X-34 staining remained unchanged in the absence of *Pld3* (Supplemental Figure 4). Thus, *Pld3* KO impacts plaque composition, shifting the pathology to a less fibrillar structure [35].

**Figure 4:**
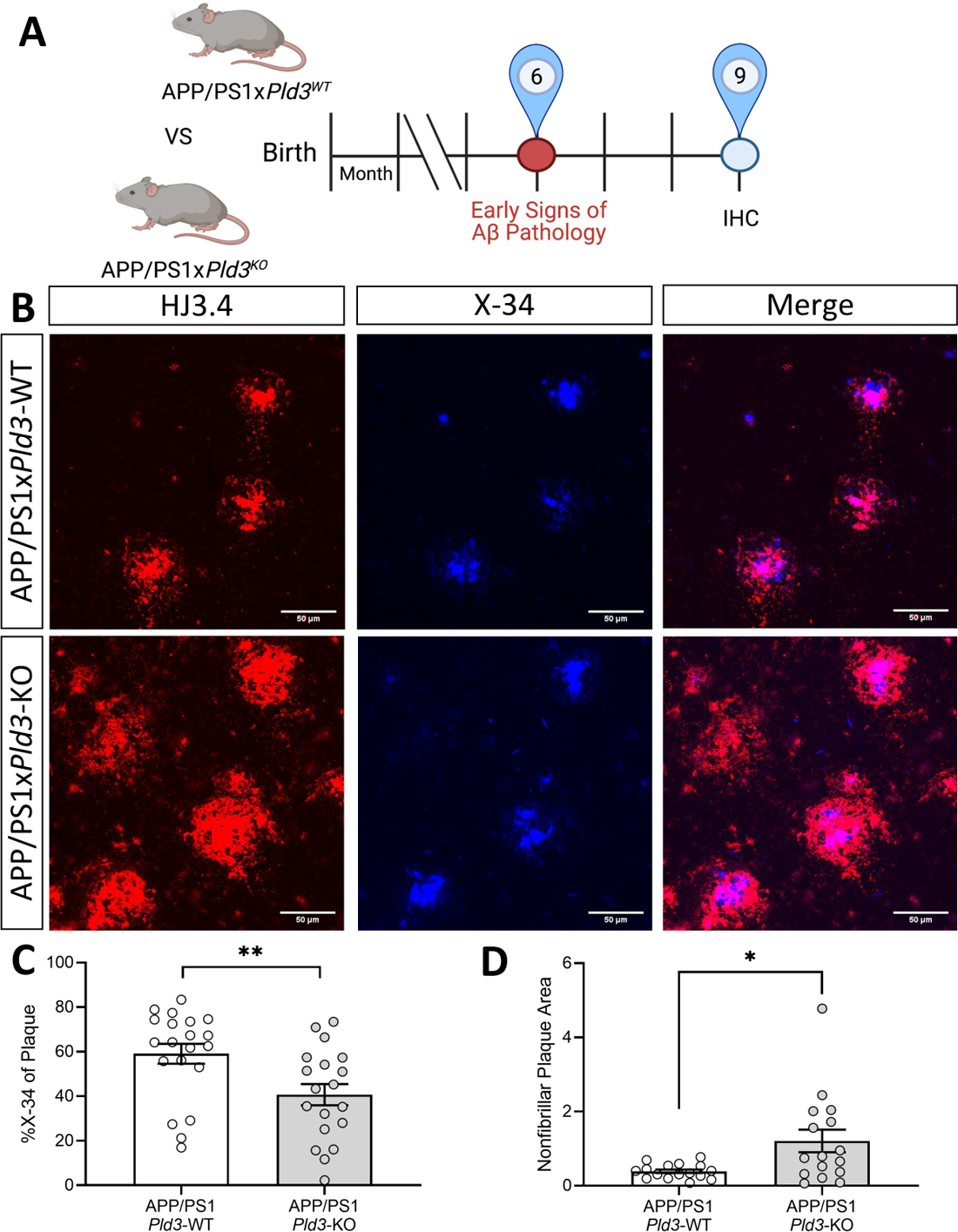
Loss of *Pld3* alters plaque composition in APP/PS1 mouse cortex. A. Experimental timeline. APP/PS1x*Pld3*-WT (n=20), APP/PS1x*Pld3*-KO (n=15). B. Representative confocal images of mouse cortex co-stained with HJ3.4 (total Aβ) and X-34 (β-sheet rich dense cores). C-D. Quantification of the plaque composition. C. Percent of X-34 within a HJ3.4-positive area. D. Area HJ3.4 outside of X-34-positive area (nonfibrillar area). The total area of X-34 normalized signal microns. Graphs represent mean ± SEM. **, p=0.0010; ***, p=0.0003. Analyzed by two-tailed Student’s *t* test with a ROUT outlier analysis (Q=1%).

### Pld3 deficiency impact microglial recruitment to amyloid plaques

The absence of *Pld3* in the APP/PS1 mice resulted in more non-fibrillar plaques. This shift in plaque composition is similar with findings from *Trem2*- and *ApoE*-deficient APP/PS1 mice [36–39]. Loss of these AD risk genes also significantly reduced microglial recruitment to the amyloid plaques [36–39]. Thus, we hypothesized that the loss of *Pld3* may alter the microglial response in APP/PS1 mice. To test this hypothesis, fixed brain tissue from the APP/PS1x*Pld3-* KO and APP/PS1x*Pld3*-WT mice were stained for total microglia (Iba1), activated microglia (CD68), and dense core Aβ plaques (X-34) (Figure 5A). The amount of activated microglia as a percentage of total microglia was similar between APP/PS1x*Pld3-*KO and APP/PS1x*Pld3*-WT mice (Figure 5D). However, APP/PS1x*Pld3*-KO mice exhibited a significant reduction in the recruitment of microglia around the X-34-positive plaques (Figure 5B). No significant change was observed in the amount of CD68-positive, activated microglia around the X-34-positive (Figure 5C and 5D). *Pld3* deficient APP/PS1 mice also exhibited a significant increase in expression of microglia genes associated with neurodegeneration including *Trem2*, *Tyrobp*, *Ctsd*, and *Cst7* (Table 1) without a corresponding change in homeostatic microglia genes (Table 1). Thus, loss of *Pld3* impacts microglia function in response to amyloid plaques.

**Figure 5:**
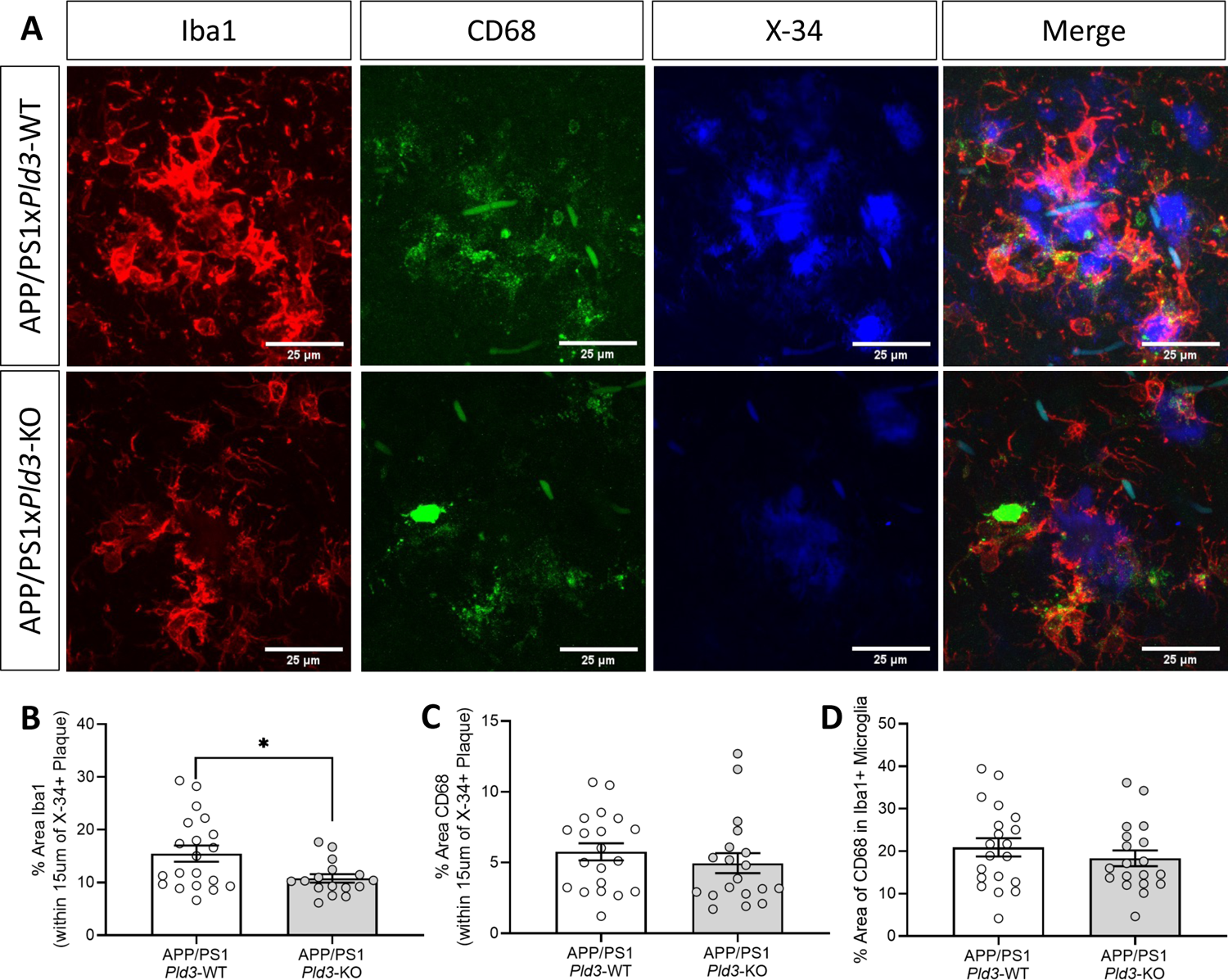
Loss of *Pld3* alters the microglial response to Aβ pathology. A. Representative images of APP/PS1x*Pld3*-WT and APP/PS1x*Pld3*-KO mice co-stained with Iba1 (total microglia), CD68 (activated microglia), and X-34 (β-sheet-rich dense cores). APP/PS1x*Pld3*-WT (n=20), APP/PS1x*Pld3*-KO (n=19). B. Quantification of Iba1 localization within 15 µm of the X-34+ dense core plaques (*, p=0.03). C. Quantification of CD68 localization within 15 µm of the dense plaques. D. Quantification of Iba1 and CD68 colocalization. Graphs represent mean ± SEM. Analyzed by two-tailed Student’s *t* test with a ROUT outlier analysis (Q=1%).

**Table 1:** Differential microglia gene expression in APP/PS1x*Pld3*-KO compared with APP/PS1x*Pld3*-WT brains

| Gene | Log2FoldChange | p-value |
| --- | --- | --- |
| *Trem2* | 0.48 | 1.07E-03 |
| *Tyrobp* | 0.57 | 1.06E-03 |
| *Ctsd* | 0.25 | 4.18E-03 |
| *Cst7* | 0.56 | 2.91E-02 |
| *Cd68* | 0.35 | 6.27E-02 |
| *Aif1* | 0.33 | 1.08E-01 |
| *Tmem119* | 0.04 | 6.49E-01 |
| *P2ry12* | 0.05 | 5.99E-01 |

### A role for PLD3 in microglia

Given the association of *Pld3* loss with altered microglia function in mouse models, we sought to determine whether *PLD3* is altered in microglia in human brains. Nuclei were isolated and sequenced from frozen AD and age-matched control brains (Figure 6A)[17, 18]. AD brains were further classified based on the presence of *TREM2* risk variants (named: TREM2). Unsupervised clustering of the brain nuclei revealed 15 cell-type specific clusters that correspond to the major cell-types found in the brain [17]. We isolated microglia from other cells and reexamined the alternative transcriptional states that we further classified into nine subclusters (Figure 6B). *PLD3* expression was significantly overexpressed in Mic.1 and reduced Mic.2 microglia subclusters compared to all microglia clusters (Figure 6B; p=2.71×10-5 and 2.27×10-6, respectively; Supplemental Table 2). In contrast, homeostatic microglia (Mic.0) did not show differential expression of *PLD3* (Figure 6B; p=0.52; Supplemental Table 2). Microglia in Mic.1 have an expression signature consistent with microglia associated with neurodegeneration (e.g. disease associated or activated response microglia) [40–42], while Mic.2 clusters are enriched among *TREM2* variant carriers and exhibit upregulated resting state microglia markers with minimal elevation of genes associated with activated microglia [17].

**Figure 6:**
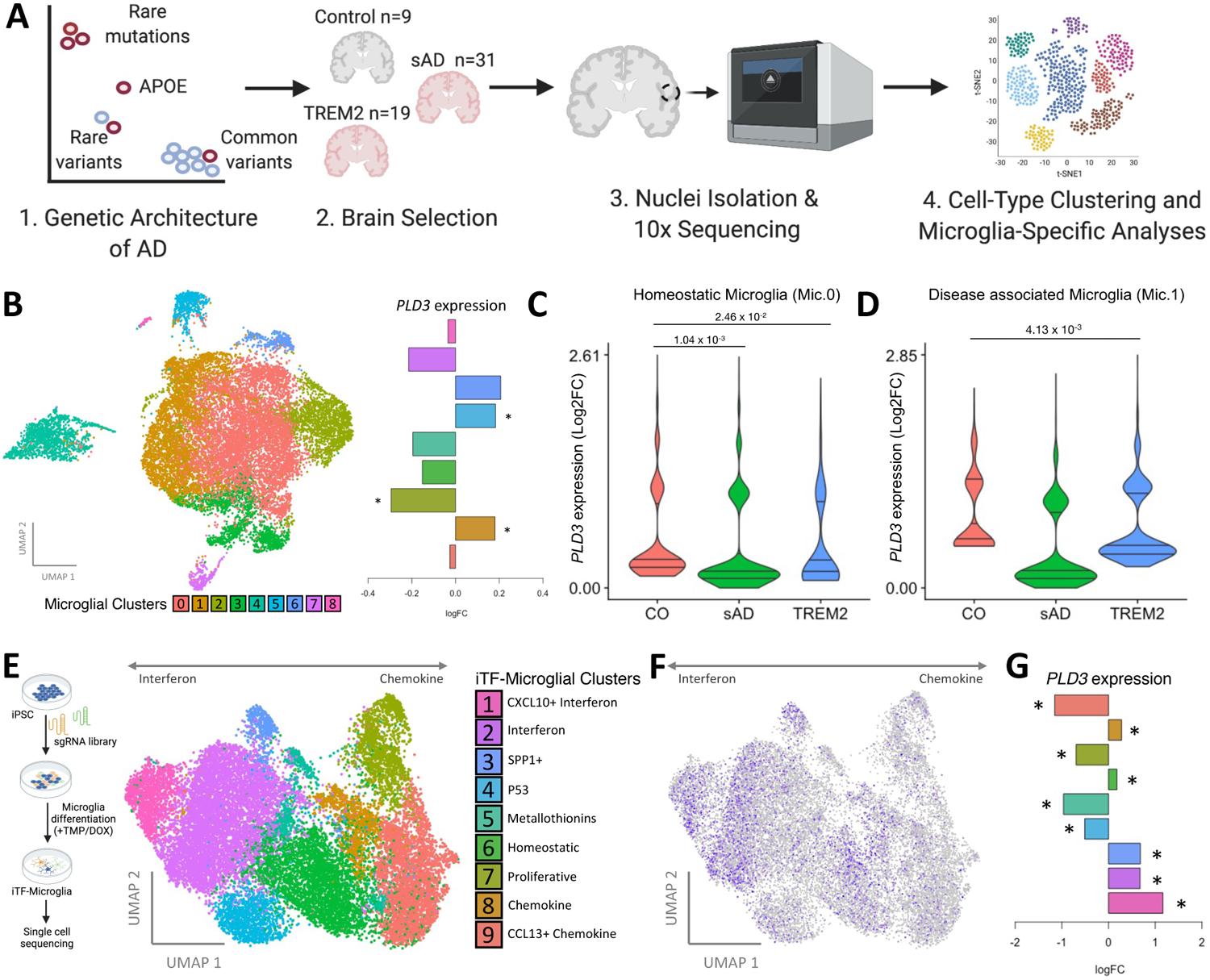
*PLD3* is enriched in specific microglia states in human brains. A. Diagram of the study design for human brain sequencing. B. UMAP plot depicting segregation of human brain microglia into nine major subclusters, left. Bar plot of the log2 fold change of *PLD3* by microglia subcluster, right. *, p<0.05. C-D. Violin plot of *PLD3* expression from control (CO), sporadic AD (sAD), and *TREM2* risk variant carriers in homeostatic (Mic.0; C) and disease associated (Mic.1; D). E-G. Single cell RNAseq data obtained iTF-Microglia CROP-seq described previously [43]. E. UMAP plot reveals 9 microglia clusters. F. Diagram of study design for iTF-Microglia, left. UMAP plot of PLD3 expression, right. Cells are colored by the *PLD3* expression levels. G. Bar plot of the log2 fold change of *PLD3* by microglia subcluster. *, p<0.05.

To understand how *PLD3* expression changes in microglia with disease, we examined control, sporadic AD and *TREM2* risk variant carriers. *PLD3* expression was significantly reduced in homeostatic microglia in sporadic AD brains and *TREM2* risk variants compared to controls (Figure 6C; p=1.04×10-3 and p=2.46×10-2, respectively; Supplemental Table 3). *PLD3* expression was further dysregulated in disease associated Mic.1 cluster in *TREM2* risk variant carriers compared with controls (Figure 6D; p=4.13×10-3; Supplemental Table 3).

To further clarify the relationship between PLD3 and microglia function, we analyzed *PLD3* expression in single cell RNAseq data obtained from human iPSC expressing inducible CRISPRi machinery that were transduced with 81 sgRNAs and differentiated into iTF-Microglia (Figure 6E) [43]. Unsupervised clustering analyses revealed nine distinct microglia subclusters (Figure 6E), representing distinct transcriptional states. Among these subclusters, *PLD3* was significantly overexpressed in clusters 1, 2, and 3 and significantly reduced in clusters 4, 5, 7, and 9 (Figure 6F and 6G; Supplemental Table 4). Clusters 1-3 correspond with interferon-induced gene activation states [43], while clusters 4-9 are enriched for genes associated with chemokine/cytokine activation states [43]. Cluster 3, where *PLD3* is significantly elevated, is enriched in *SPP1* expression, a marker of disease associated microglia [43]. Additionally, cluster 7, where *PLD3* is significantly reduced, is enriched in markers of microglia proliferation [43]. Together, these data support a role for PLD3 in microglia activation in health and disease.

## Discussion

In this study, we sought to understand the contribution of PLD3 to pathways that promote AD pathology. We demonstrate that the AD risk variant, *PLD3* p.A442A, is sufficient to alter *PLD3* splicing and Aβ levels in iPSC-derived neurons in a manner consistent with similar findings in AD brains [4]. Additionally, we describe a role for PLD3 in LOAD, whereby modifying *PLD3* expression in APP/PS1 mice is sufficient to regulate Aβ turnover in the ISF. The observed reduced ISF Aβ turnover, in turn, leads to a change in amyloid plaques in aged animals. We observed that loss of *Pld3* in APP/PS1 mice results in a shift in plaque composition to a more nonfibrillar structure. This altered plaque composition is accompanied by impaired microglial recruitment to the plaques, consistent with prior reports from *Trem2* deficient mice. In human brains, *PLD3* is enriched in disease associated microglia and expression is altered in AD brains. Together, these results suggest that *PLD3* plays cell-autonomous and non-cell autonomous roles in AD pathogenesis.

Deciphering the contribution of risk variants and pathogenic mutations to AD pathogenesis has led to groundbreaking discoveries of Aβ metabolism, synaptic function, and immune function to AD and revealed novel therapeutic targets [44]. Emerging sequencing technologies in increasingly larger cohorts have revealed the contribution of rare variants to AD risk [44, 45]. Nevertheless, resolving the contribution of rare variants to disease can be challenging when relying on association studies and autopsy brain tissue that captures a snapshot of disease.

Here, patient-derived cell culture models represent a tractable, human platform that recapitulates disease-specific phenotypes and when coupled with genome engineering, allows for the study of genotype x phenotype relationships. This study demonstrates that iPSC-derived neurons are highly informative and recapitulate early pathogenic events in AD.

In this study, we used genome editing technology to molecularly pinpoint the contribution of the A allele to PLD3 and AD-related phenotypes. While we cannot exclude the possibility that genomic factors beyond PLD3 p.A442A contribute to the risk profile in the iPSC donor line used in this study, we can attribute the defect in PLD3 splicing and the increase in Aβ levels to this synonymous variant. The increase in both Aβ42 and Aβ40 is consistent with the effects of other known pathogenic mutations, including APP KM670/671NL [46, 47]. The absence of an effect of *PLD3* p.A442A on the Aβ42/40 ratio suggests that Aβ recycling and trafficking. This is consistent with recently reported functions of PLD3 as a type II membrane protein functioning in endosomes and lysosomes, the primary site of APP cleavage [48, 49]. Together, these human stem cell findings suggest a role for PLD3 p.A442A in altering APP/Aβ recycling and trafficking in a manner that elevates total Aβ levels. PLD3 p.A442A was predicted to disrupt a splicing enhancer-binding site [4]. We observe defective *PLD3* splicing in iPSC-derived neurons from a *PLD3* p.A442A carrier, which replicates the observations in brains from *PLD3* p.A442A carriers [4]. We go on to demonstrate that correcting the risk allele with CRISPR/Cas9 is sufficient to restore the splicing defect. The functional impact of distinct PLD3 isoforms remains unknown; however, as the functional roles of PLD3 are resolved, this will be an important area to explore.

We show that PLD3 is a major regulator of ISF Aβ turnover *in vivo*. Hippocampal reduction of endogenous *Pld3* in adulthood via AAV8-mediated knockdown or global knockout of *Pld3* in the background of APP/PS1 mice resulted in a strong increase in ISF Aβ half-life, suggesting that Aβ is turned over more slowly in the absence of *Pld3*. Aβ clearance mechanisms have been proposed to drive LOAD [33]. Following secretion from presynaptic neurons, Aβ is either taken up and degraded in the lysosomes of post-synaptic neurons, taken up and degraded in the lysosomes of glial cells, degraded by extracellular proteases, is transported to CSF, or transcytosed across the blood-brain barrier (BBB). Amyloid accumulation occurs in an Aβ concentration-dependent manner [34]; thus, dysregulation of ISF Aβ clearance drives amyloid accumulation and AD pathogenesis. Neurons from *Pld3* KO mice exhibit lysosomes with increased density and size [31]. PLD3 is enriched in lysosomes surrounding amyloid plaques in human AD brains and mouse models of amyloid accumulation [9]. Thus, *Pld3* may regulate ISF Aβ through lysosome-mediated clearance mechanisms in neurons or other glial cells.

The absence of *Pld3* in APP/PS1 mice led to a shift in the composition of amyloid plaques to being more diffuse and less fibrillar. While amyloid plaques may adopt a series of morphologies and architecture, a major structure is characterized by a dense “fibrillar” core of Aβ surrounded by more diffuse “non-fibrillar” of Aβ deposits [50]. Non-fibrillar Aβ is proposed to contribute to its higher toxicity, possible because either its structure is more toxic or they serve as a reservoir for more diffusible Aβ oligomers [35, 51]. Thus, a PLD3-mediated shift to more non-fibrillar plaques is consistent with more toxic effects given the same amount of overall Aβ deposition. Silencing AD risk genes, including *Trem2* and *ApoE,* result in a similar shift of plaque composition shift in APP/PS1 mice [36, 37, 39]. Thus, modeling the reduction of *PLD3* observed in *PLD3* p.A442A carriers, and LOAD brains in an animal model of amyloid accumulation leads to a phenotype consistent with a gene that exacerbates disease.

In prior studies where gene silencing in amyloid mice led to a change in plaque formation and composition, a role for an altered microglial response to amyloid plaques was implicated [36, 37, 39]. APP/PS1x*Pld3*-KO and APP/PS1x*Pld3*-WT mice exhibited a similar abundance of Iba1 positive microglia. Yet, recruitment of Iba1-positive microglia to X-34-positive plaques was significantly reduced in APP/PS1x*Pld3*-KO mice compared to APP/PS1x*Pld3*-WT mice. This could suggest a role for PLD3 in recruiting microglia to surround and alter Aβ structure and limit Aβ-induced toxicity similar to mechanisms described for TREM2 [36]. In mice, *Pld3* mRNA is expressed in microglia [52]. Microglia isolated from amyloid mouse models (APP^NL-F-G^) reveal an activation state enriched for MHC class II, tissue repair genes, and enrichment of AD risk genes, including *Pld3* [42]. Thus, loss of *Pld3* in the global knockout may impact the molecular identity of microglia, which impairs the recruitment and responsiveness of the glia to plaques.

In human microglia, PLD3 plays a role in microglia that is disrupted in AD. Microglia maintain distinct transcriptional states that likely reflect functional changes due to environmental stimuli [53]. We demonstrate that *PLD3* expression is enriched in disease associated microglia (Brain Mic.1 and iTF-Microglia cluster 3) and depleted in population of microglia that are found in *TREM2* risk variant carriers (Brain Mic.2). Mic.2 reflect a dampened activation state, distinct from homeostatic microglia (Mic.0) with an upregulation of resting state microglia markers (*TMEM119, P2RY13, MED12L*) and modest elevation of activated markers (*ABCA1, C5AR1,* and *CD83*) [17]. This finding along with the observation that *PLD3* expression is reduced in disease associated microglia (Mic.1) in *TREM2* risk variant carriers suggests a potential interaction between these AD risk genes. These results also support the parallels between our mouse model findings and those in *Trem2* deficient mice. In addition to association with disease associated microglia, PLD3 and TREM2 have also been implicated in lysosomal function [9, 31, 38, 48, 49].

Overall, we observed a modest impact on amyloid plaque pathology in APP/PS1x*Pld3*-KO mice. The modest impact is highly consistent with PLD3 as a disease modifier rather than a fully penetrant, causative mutation. Alternatively, this could reflect redundant mechanisms for mouse Pld3.

Here, we demonstrate a therapeutic potential for PLD3. *hPLD3* overexpression by AAV8 in APP/PS1 mice resulted in a significant decrease in ISF Aβ levels and accelerated Aβ turnover. *PLD3* levels are significantly reduced in AD brains, and *PLD3* expression is positively correlated with cognition in humans and mouse models [9]. Thus, by promoting Aβ turnover and facilitating the microglial response to amyloid plaques, *PLD3* occupies a crucial role in brain health.

## Data Availability

All data produced in the present study are available upon reasonable request to the authors. Data can be publicly accessed at http://ngi.pub/SNARE.

## Acknowledgments

This work was funded by BrightFocus Foundation (CMK), the Alzheimer’s Association (CMK), NIH U01 AG052411 (AG), R01 AG062359 (CMK), P50 AG005681 (JM, CMK), R56AG067764 (CMK, OH, CC), U01 AG072464 (ST, CMK, OH, MK), UL1 TR002345, NS090934 (DMH), AG047644 (DMH), NS094692 (JML), AG062359 (MK). This work was supported by access to equipment made possible by the Hope Center for Neurological Disorders and the Departments of Neurology and Psychiatry at Washington University School of Medicine. We thank the Washington University in St Louis Mouse Genetics Core for assistance in generating the Pld3 KO mice. The gRNAs were generated by the Genome Engineering and iPSC Center (GEiC) at the Washington University in St. Louis. We thank the Hope Center Viral Vector Core for generating the AAV8 particles and the Hope Center Animal Surgery Core for performing the AAV8 injections. Confocal was generated on a Zeiss LSM 880 Airyscan Confocal Microscope, which was purchased with support from the Office of Research Infrastructure Programs (ORIP), a part of the NIH Office of the Director under grant OD021629. The recruitment and clinical characterization of research participants at Washington University were supported by NIH P50 AG05681, P01 AG03991, and P01 AG026276.

## Supplemental Figure Legends

**Supplemental Figure 1:**
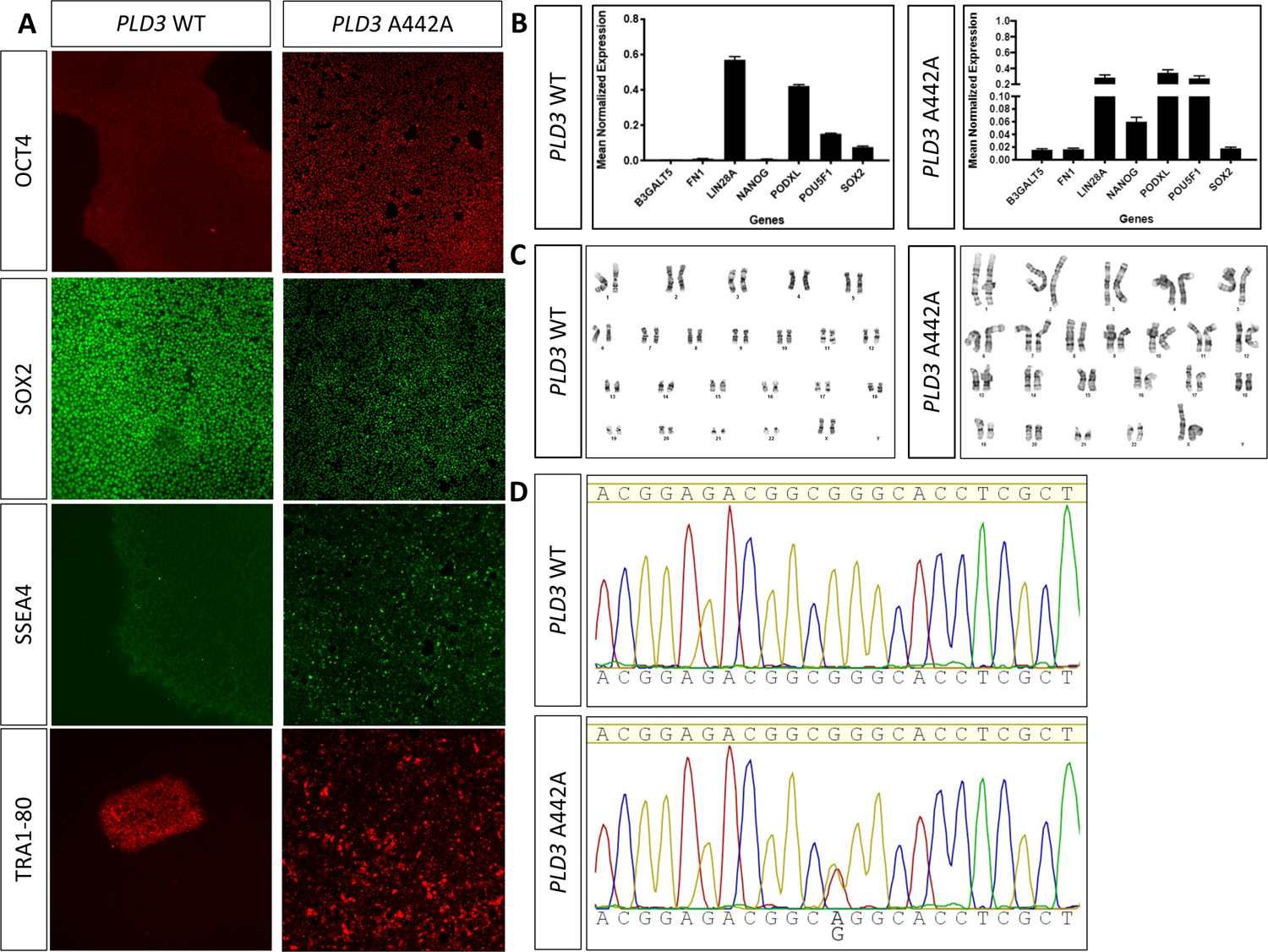
Characterization of the *PLD3* p.A442A iPSC-derived neurons. A. Representative images of *PLD3* p.A442A and the corrected WT iPSCs stained for NANOG, OCT4, SOX2, SSEA4, and TRA1-80. B. qPCR from known markers of pluripotency. C. Karyotype. D. Sanger sequencing.

**Supplemental Figure 2:**
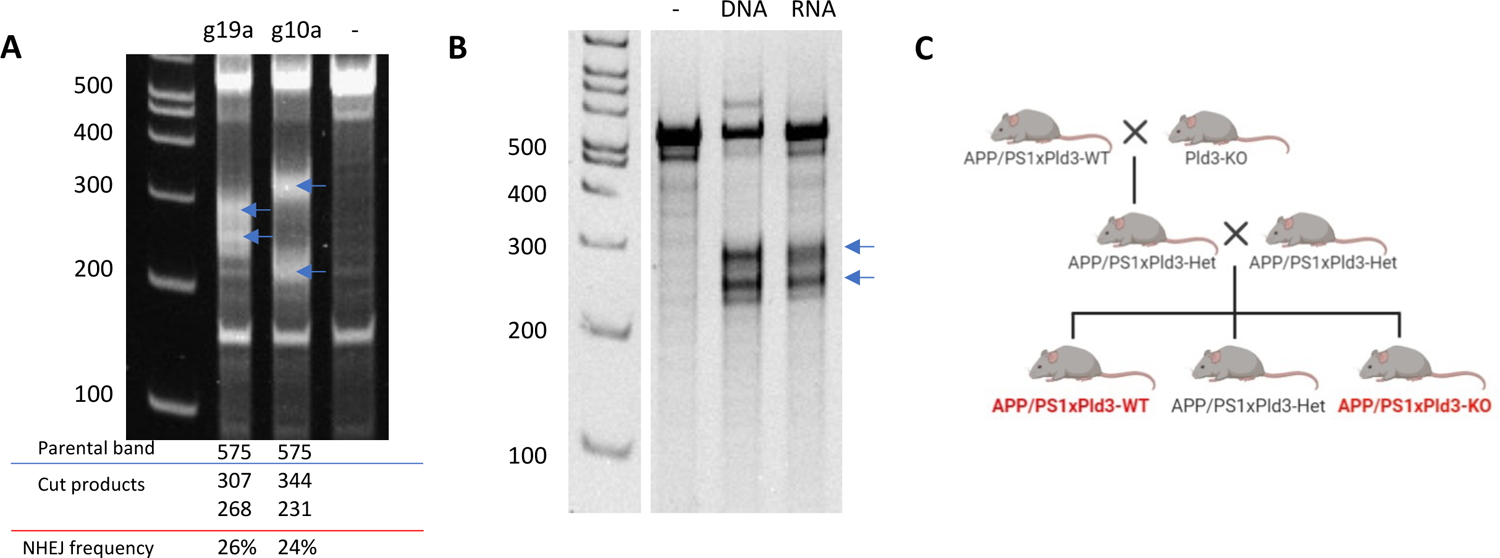
Generation of a *Pld3*-deficient amyloid mouse model. A. Mismatch detection assay. B. RNA activity validation. C. Breeding scheme for the *Pld3*-deficient mouse with APP/PS1 mutant mice to develop a transgenic APP/PS1x*Pld3*-KO mouse line along with APP/PS1x*Pld3*-WT littermate controls.

**Supplemental Figure 3:**
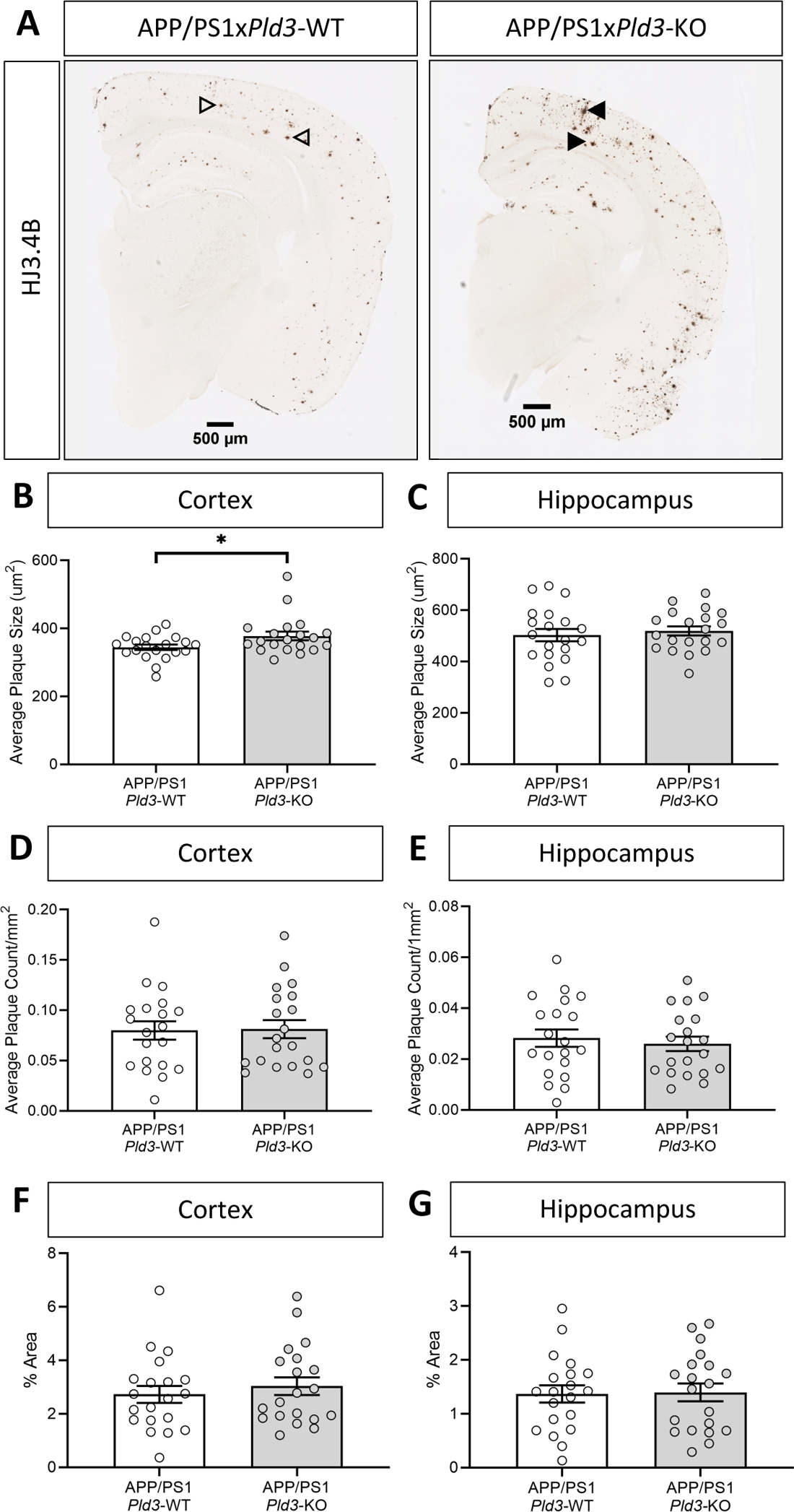
Loss of *Pld3* significantly increases plaque size without changing plaque area. A. Representative images of mice brain cross-sections stained for total Aβ (HJ3.4) with arrows specifying plaques (open arrows for WT; closed arrows for KO). APP/PS1x*Pld3*-WT (n=20), APP/PS1x*Pld3*-KO (n=20). B-C. Quantification of average plaque size in the cortex (B) and hippocampus (C). D-E. Quantification of average plaque count per mm^2^ in the cortex (D) and the hippocampus (E). F-G. Quantification of the plaque burden by the percentage of the total area for the cortex (F) and the hippocampus (G). Graphs represent mean ± SEM. *, p>0.05.

**Supplemental Figure 4:**
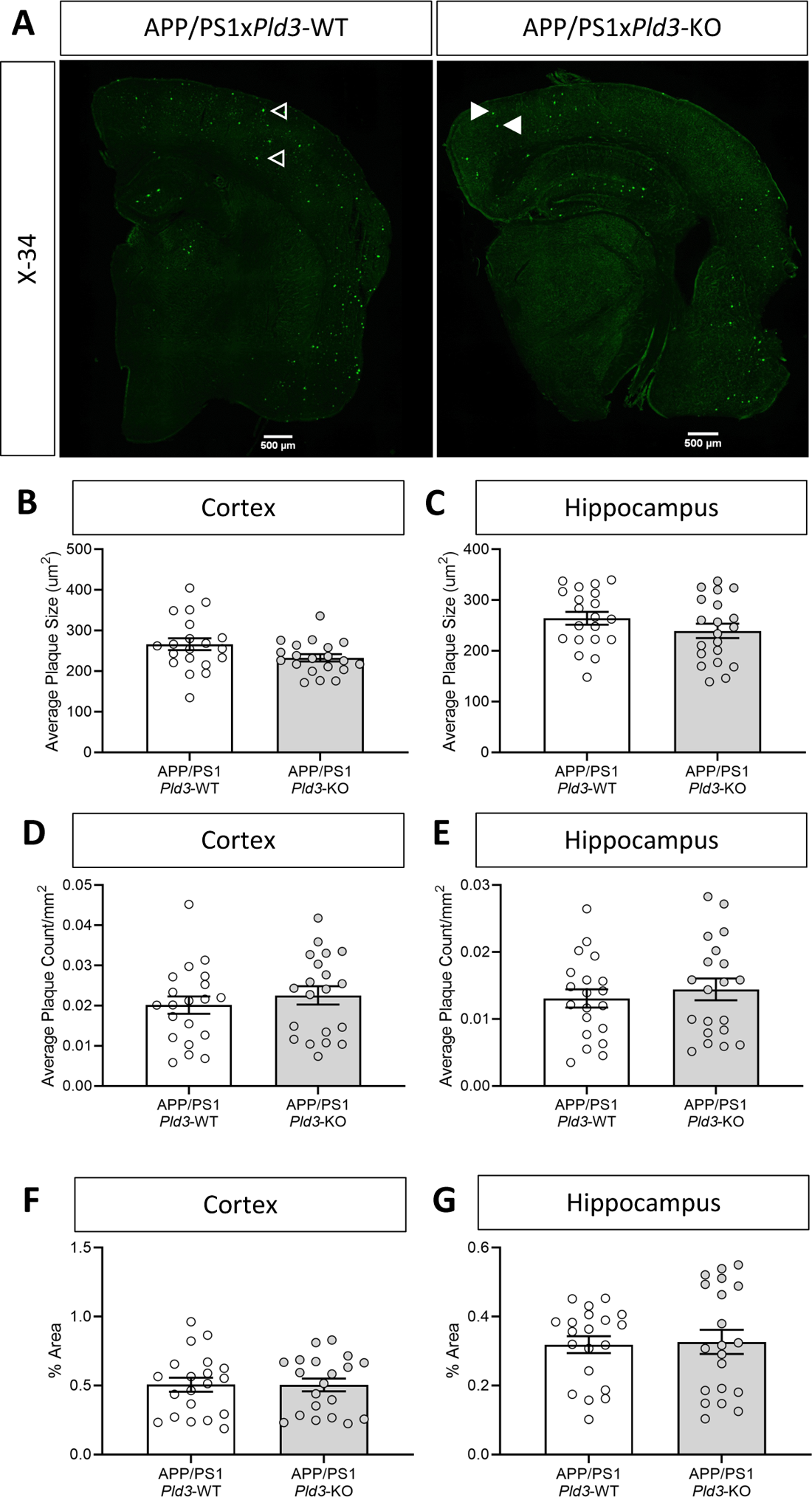
Loss of *Pld3* does not change dense core plaques. A. Representative images of mice brain cross sections stained with X-34 with arrows specifying plaques (open arrows for WT; closed arrows for KO). APP/PS1x*Pld3*-WT (n=20), APP/PS1x*Pld3*-KO (n=20). B-C. Quantification of average plaque size in the cortex (B) and hippocampus (C). D-E. Quantification of average plaque count per mm^2^ in the cortex (D) and hippocampus (E). F-G. The quantification of the plaque burden by the percentage of the total area for the cortex (F) and hippocampus (G). Graphs represent mean ± SEM.

## Supplemental Tables

**Supplemental Table 1:** Human brain demographics for snRNAseq

| Supplemental Table 1: Human brain demographics for snRNAseq |  |  |  |  |  |
| --- | --- | --- | --- | --- | --- |
|  | Total Number | Sex (% male) | Mean Age at Death (years) | APOE4 | Mean Postmortem Interval (years) |
| Controls | 9 | 33% | 90 | 11% | 10.9 |
| sAD | 31 | 45% | 81 | 54% | 11.9 |

**Supplemental Table 2:** PLD3 expression in human brain microglia

| Supplemental Table 2: PLD3 expression in human brain microglia |  |  |  |  |
| --- | --- | --- | --- | --- |
| Cluster Name | LogFC | z-score | p-value | BH corrected p-value |
| Mic.0 | -0.0262225 | -0.6410652 | 5.21E-01 | 7.32E-01 |
| Mic.1 | 0.1800801 | 4.19659881 | 2.71E-05 | 2.86E-04 |
| Mic.2 | -0.2949385 | -4.7275062 | 2.27E-06 | 1.26E-04 |
| Mic.3 | -0.1519921 | -1.274392 | 2.03E-01 | 5.14E-01 |
| Mic.4 | -0.1953743 | -1.718988 | 8.56E-02 | 2.51E-01 |
| Mic.5 | 0.1829342 | 1.98158159 | 4.75E-02 | 1.75E-01 |
| Mic.6 | 0.20610645 | 1.89898926 | 5.76E-02 | 1.96E-01 |
| Mic.7 | -0.2146737 | -1.3839443 | 1.66E-01 | 4.50E-01 |
| Mic.8 | -0.0345632 | -0.1138777 | 9.09E-01 | 9.99E-01 |

**Supplemental Table 3:** PLD3 expression within microglia subclusters by disease status

|  |  |  |  |  |  |
| --- | --- | --- | --- | --- | --- |
| Comparison | LogFC | z-score | p-value | BH corrected p-value | Subcluster |
| sAD vs CO | -0.4017806 | -3.2799941 | 1.04E-03 | 1.43E-01 | Mic.0 |
| TREM2 vs CO | -0.2766865 | -2.247029 | 2.46E-02 | 3.91E-01 | Mic.0 |
| sAD vs CO | -0.3455155 | -1.8251283 | 6.80E-02 | 1.00E+00 | Mic.1 |
| TREM2 vs CO | -0.4217827 | -2.8679334 | 4.13E-03 | 3.21E-01 | Mic.1 |

**Supplemental Table 4:**
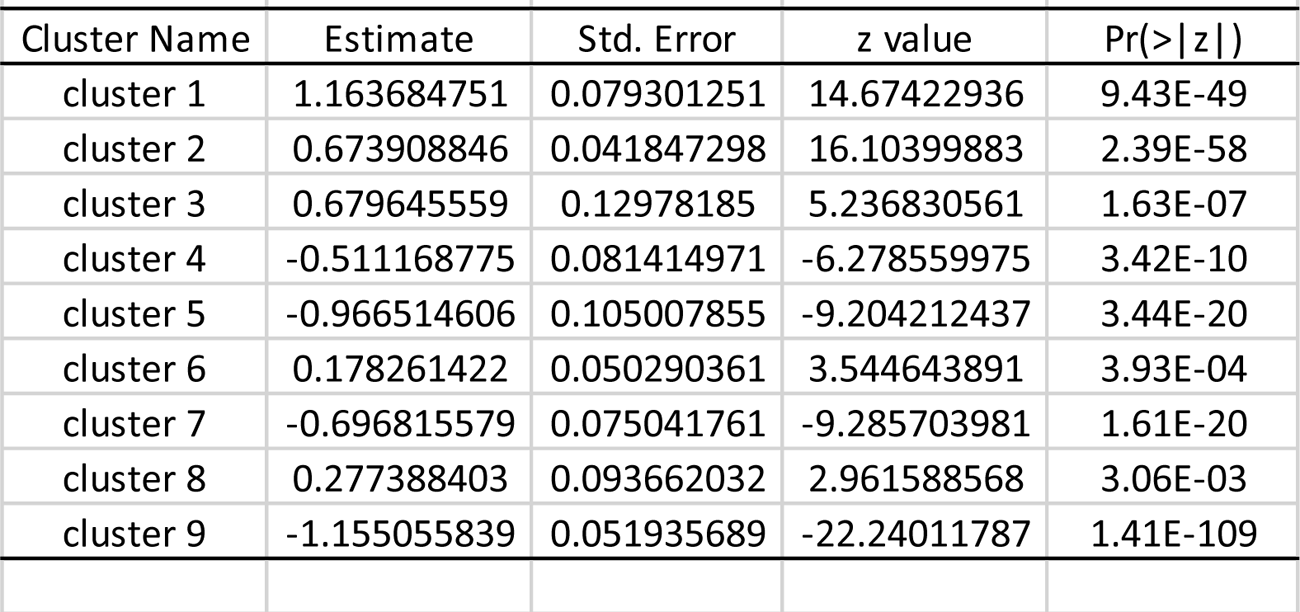
PLD3 expression in iTF-Microglia CROP-Seq

## References

1. J. Hardy, D.J. Selkoe, The amyloid hypothesis of Alzheimer’s disease: progress and problems on the road to therapeutics, Science 297(5580) (2002) 353-6.

2. C.M. Karch, A.M. Goate, Alzheimer’s disease risk genes and mechanisms of disease pathogenesis, Biol Psychiatry 77(1) (2015) 43–51.

3. S.M. Neuner, J. Tcw, A.M. Goate, Genetic architecture of Alzheimer’s disease, Neurobiol Dis 143 (2020) 104976.

4. C. Cruchaga, C.M. Karch, S.C. Jin, B.A. Benitez, Y. Cai, R. Guerreiro, O. Harari, J. Norton, J. Budde, S. Bertelsen, A.T. Jeng, B. Cooper, T. Skorupa, D. Carrell, D. Levitch, S. Hsu, J. Choi, M. Ryten, C. Sassi, J. Bras, R.J. Gibbs, D.G. Hernandez, M.K. Lupton, J. Powell, P. Forabosco, P.G. Ridge, C.D. Corcoran, J.T. Tschanz, M.C. Norton, R.G. Munger, C. Schmutz, M. Leary, F.Y. Demirci, M.N. Bamne, X. Wang, O.L. Lopez, M. Ganguli, C. Medway, J. Turton, J. Lord, A. Braae, I. Barber, K. Brown, U.K.C. Alzheimer’s Research, P. Pastor, O. Lorenzo-Betancor, Z. Brkanac, E. Scott, E. Topol, K. Morgan, E. Rogaeva, A. Singleton, J. Hardy, M.I. Kamboh, P.S. George-Hyslop, N. Cairns, J.C. Morris, J.S.K. Kauwe, A.M. Goate, Rare coding variants in the phospholipase D3 gene confer risk for Alzheimer’s disease, Nature 505(7484) (2014) 550-554.

5. M. Osisami, W. Ali, M.A. Frohman, A role for phospholipase D3 in myotube formation, PloS one 7(3) (2012) e33341.

6. A. Munck, C. Bohm, N.M. Seibel, Z. Hashemol Hosseini, W. Hampe, Hu-K4 is a ubiquitously expressed type 2 transmembrane protein associated with the endoplasmic reticulum, The FEBS journal 272(7) (2005) 1718–26.

7. K.M. Pedersen, B. Finsen, J.E. Celis, N.A. Jensen, Expression of a novel murine phospholipase D homolog coincides with late neuronal development in the forebrain, The Journal of biological chemistry 273(47) (1998) 31494–504.

8. J. Satoh, Y. Kino, Y. Yamamoto, N. Kawana, T. Ishida, Y. Saito, K. Arima, PLD3 is accumulated on neuritic plaques in Alzheimer’s disease brains, Alzheimers Res Ther 6(9) (2014) 70.

9. A.G. Nackenoff, T.J. Hohman, S.M. Neuner, C.S. Akers, N.C. Weitzel, A. Shostak, S.M. Ferguson, B. Mobley, D.A. Bennett, J.A. Schneider, A.L. Jefferson, C.C. Kaczorowski, M.S. Schrag, PLD3 is a neuronal lysosomal phospholipase D associated with beta-amyloid plaques and cognitive function in Alzheimer’s disease, PLoS Genet 17(4) (2021) e1009406.

10. C. Wang, H.F. Wang, M.S. Tan, Y. Liu, T. Jiang, D.Q. Zhang, L. Tan, J.T. Yu, I. Alzheimer’s Disease Neuroimaging, Impact of Common Variations in PLD3 on Neuroimaging Phenotypes in Non-demented Elders, Mol Neurobiol 53(7) (2016) 4343–51.

11. K. Takahashi, S. Yamanaka, Induction of pluripotent stem cells from mouse embryonic and adult fibroblast cultures by defined factors, Cell 126(4) (2006) 663–76.

12. H. Ban, N. Nishishita, N. Fusaki, T. Tabata, K. Saeki, M. Shikamura, N. Takada, M. Inoue, M. Hasegawa, S. Kawamata, S. Nishikawa, Efficient generation of transgene-free human induced pluripotent stem cells (iPSCs) by temperature-sensitive Sendai virus vectors, Proceedings of the National Academy of Sciences of the United States of America 108(34) (2011) 14234–9.

13. C.M. Karch, A.W. Kao, A. Karydas, K. Onanuga, R. Martinez, A. Argouarch, C. Wang, C. Huang, P.D. Sohn, K.R. Bowles, S. Spina, M.C. Silva, J.A. Marsh, S. Hsu, D.A. Pugh, N. Ghoshal, J. Norton, Y. Huang, S.E. Lee, W.W. Seeley, P. Theofilas, L.T. Grinberg, F. Moreno, K. McIlroy, B.F. Boeve, N.J. Cairns, J.F. Crary, S.J. Haggarty, J.K. Ichida, K.S. Kosik, B.L. Miller, L. Gan, A.M. Goate, S. Temple, G. Tau Consortium Stem Cell, A Comprehensive Resource for Induced Pluripotent Stem Cells from Patients with Primary Tauopathies, Stem Cell Reports 13(5) (2019) 939–955.

14. S.W. Cho, S. Kim, J.M. Kim, J.S. Kim, Targeted genome engineering in human cells with the Cas9 RNA-guided endonuclease, Nat Biotechnol 31(3) (2013) 230–2.

15. Z. Li, J.L. Del-Aguila, U. Dube, J. Budde, R. Martinez, K. Black, Q. Xiao, N.J. Cairns, N. Dominantly Inherited Alzheimer, J.D. Dougherty, J.M. Lee, J.C. Morris, R.J. Bateman, C.M. Karch, C. Cruchaga, O. Harari, Genetic variants associated with Alzheimer’s disease confer different cerebral cortex cell-type population structure, Genome Med 10(1) (2018) 43.

16. M. Allen, M.M. Carrasquillo, C. Funk, B.D. Heavner, F. Zou, C.S. Younkin, J.D. Burgess, H.-S. Chai, J. Crook, J.A. Eddy, H. Li, B. Logsdon, M.A. Peters, K.K. Dang, X. Wang, D. Serie, C. Wang, T. Nguyen, S. Lincoln, K. Malphrus, G. Bisceglio, M. Li, T.E. Golde, L.M. Mangravite, Y. Asmann, N.D. Price, R.C. Petersen, N.R. Graff-Radford, D.W. Dickson, S.G. Younkin, N. Ertekin-Taner, Human whole genome genotype and transcriptome data for Alzheimer’s and other neurodegenerative diseases, Scientific Data 3(1) (2016) 160089.

17. L. Brase, S.-F. You, J. del Aguila, Y. Dai, B.C. Novotny, C. Soriano-Tarraga, T. Dykstra, M.V. Fernandez, J.P. Budde, K. Bergmann, J.C. Morris, R.J. Bateman, R.J. Perrin, E. McDade, C. Xiong, A. Goate, M. Farlow, J.P. Chhatwal, P. Schofield, H. Chui, D.I.A. Network, G.T. Sutherland, J. Kipnis, C.M. Karch, B.A. Benitez, C. Cruchaga, O. Harari, A landscape of the genetic and cellular heterogeneity in Alzheimer disease, medRxiv (2021) 2021.11.30.21267072.

18. S. Da Mesquita, Z. Papadopoulos, T. Dykstra, L. Brase, F.G. Farias, M. Wall, H. Jiang, C.D. Kodira, K.A. de Lima, J. Herz, A. Louveau, D.H. Goldman, A.F. Salvador, S. Onengut-Gumuscu, E. Farber, N. Dabhi, T. Kennedy, M.G. Milam, W. Baker, I. Smirnov, S.S. Rich, N. Dominantly Inherited Alzheimer, B.A. Benitez, C.M. Karch, R.J. Perrin, M. Farlow, J.P. Chhatwal, D.M. Holtzman, C. Cruchaga, O. Harari, J. Kipnis, Meningeal lymphatics affect microglia responses and anti-Abeta immunotherapy, Nature 593(7858) (2021) 255-260.

19. J.L. Jankowsky, D.J. Fadale, J. Anderson, G.M. Xu, V. Gonzales, N.A. Jenkins, N.G. Copeland, M.K. Lee, L.H. Younkin, S.L. Wagner, S.G. Younkin, D.R. Borchelt, Mutant presenilins specifically elevate the levels of the 42 residue beta-amyloid peptide in vivo: evidence for augmentation of a 42-specific gamma secretase, Hum Mol Genet 13(2) (2004) 159–70.

20. Q. Xiao, P. Yan, X. Ma, H. Liu, R. Perez, A. Zhu, E. Gonzales, D.L. Tripoli, L. Czerniewski, A. Ballabio, J.R. Cirrito, A. Diwan, J.M. Lee, Neuronal-Targeted TFEB Accelerates Lysosomal Degradation of APP, Reducing Abeta Generation and Amyloid Plaque Pathogenesis, J Neurosci 35(35) (2015) 12137–51.

21. Q. Xiao, S.C. Gil, P. Yan, Y. Wang, S. Han, E. Gonzales, R. Perez, J.R. Cirrito, J.M. Lee, Role of phosphatidylinositol clathrin assembly lymphoid-myeloid leukemia (PICALM) in intracellular amyloid precursor protein (APP) processing and amyloid plaque pathogenesis, The Journal of biological chemistry 287(25) (2012) 21279–89.

22. P. Yan, A.W. Bero, J.R. Cirrito, Q. Xiao, X. Hu, Y. Wang, E. Gonzales, D.M. Holtzman, J.-M. Lee, Characterizing the appearance and growth of amyloid plaques in APP/PS1 mice, J Neurosci 29(34) (2009) 10706–14.

23. D.R. Borchelt, T. Ratovitski, J. van Lare, M.K. Lee, V. Gonzales, N.A. Jenkins, N.G. Copeland, D.L. Price, S.S. Sisodia, Accelerated amyloid deposition in the brains of transgenic mice coexpressing mutant presenilin 1 and amyloid precursor proteins, Neuron 19(4) (1997) 939–45.

24. Q. Xiao, P. Yan, X. Ma, H. Liu, R. Perez, A. Zhu, E. Gonzales, J.M. Burchett, D.R. Schuler, J.R. Cirrito, A. Diwan, J.M. Lee, Enhancing astrocytic lysosome biogenesis facilitates Abeta clearance and attenuates amyloid plaque pathogenesis, J Neurosci 34(29) (2014) 9607–20.

25. J.R. Cirrito, P.C. May, M.A. O’Dell, J.W. Taylor, M. Parsadanian, J.W. Cramer, J.E. Audia, J.S. Nissen, K.R. Bales, S.M. Paul, R.B. DeMattos, D.M. Holtzman, In vivo assessment of brain interstitial fluid with microdialysis reveals plaque-associated changes in amyloid-beta metabolism and half-life, The Journal of neuroscience: the official journal of the Society for Neuroscience 23(26) (2003) 8844–53.

26. J.R. Cirrito, K.A. Yamada, M.B. Finn, R.S. Sloviter, K.R. Bales, P.C. May, D.D. Schoepp, S.M. Paul, S. Mennerick, D.M. Holtzman, Synaptic activity regulates interstitial fluid amyloid-beta levels in vivo, Neuron 48(6) (2005) 913–22.

27. J.C. Hettinger, H. Lee, G. Bu, D.M. Holtzman, J.R. Cirrito, AMPA-ergic regulation of amyloid-beta levels in an Alzheimer’s disease mouse model, Mol Neurodegener 13(1) (2018) 22.

28. W. Farris, S.G. Schutz, J.R. Cirrito, G.M. Shankar, X. Sun, A. George, M.A. Leissring, D.M. Walsh, W.Q. Qiu, D.M. Holtzman, D.J. Selkoe, Loss of neprilysin function promotes amyloid plaque formation and causes cerebral amyloid angiopathy, The American journal of pathology 171(1) (2007) 241–51.

29. J.H. Roh, Y. Huang, A.W. Bero, T. Kasten, F.R. Stewart, R.J. Bateman, D.M. Holtzman, Disruption of the sleep-wake cycle and diurnal fluctuation of beta-amyloid in mice with Alzheimer’s disease pathology, Sci Transl Med 4(150) (2012) 150ra122.

30. H. Braak, E. Braak, Neuropathological stageing of Alzheimer-related changes, Acta Neuropathol 82(4) (1991) 239–59.

31. P. Fazzari, K. Horre, A.M. Arranz, C.S. Frigerio, T. Saito, T.C. Saido, B. De Strooper, PLD3 gene and processing of APP, Nature 541(7638) (2017) E1-E2.

32. J.M. Tarasoff-Conway, R.O. Carare, R.S. Osorio, L. Glodzik, T. Butler, E. Fieremans, L. Axel, H. Rusinek, C. Nicholson, B.V. Zlokovic, B. Frangione, K. Blennow, J. Menard, H. Zetterberg, T. Wisniewski, M.J. de Leon, Clearance systems in the brain-implications for Alzheimer disease, Nat Rev Neurol 11(8) (2015) 457–70.

33. K.G. Mawuenyega, W. Sigurdson, V. Ovod, L. Munsell, T. Kasten, J.C. Morris, K.E. Yarasheski, R.J. Bateman, Decreased clearance of CNS beta-amyloid in Alzheimer’s disease, Science 330(6012) (2010) 1774.

34. A.W. Bero, P. Yan, J.H. Roh, J.R. Cirrito, F.R. Stewart, M.E. Raichle, J.M. Lee, D.M. Holtzman, Neuronal activity regulates the regional vulnerability to amyloid-beta deposition, Nat Neurosci 14(6) (2011) 750–6.

35. C.M. Vander Zanden, L. Wampler, I. Bowers, E.B. Watkins, J. Majewski, E.Y. Chi, Fibrillar and Nonfibrillar Amyloid Beta Structures Drive Two Modes of Membrane-Mediated Toxicity, Langmuir 35(48) (2019) 16024–16036.

36. Y. Wang, T.K. Ulland, J.D. Ulrich, W. Song, J.A. Tzaferis, J.T. Hole, P. Yuan, T.E. Mahan, Y. Shi, S. Gilfillan, M. Cella, J. Grutzendler, R.B. DeMattos, J.R. Cirrito, D.M. Holtzman, M. Colonna, TREM2-mediated early microglial response limits diffusion and toxicity of amyloid plaques, J Exp Med 213(5) (2016) 667–75.

37. J.D. Ulrich, T.K. Ulland, T.E. Mahan, S. Nystrom, K.P. Nilsson, W.M. Song, Y. Zhou, M. Reinartz, S. Choi, H. Jiang, F.R. Stewart, E. Anderson, Y. Wang, M. Colonna, D.M. Holtzman, ApoE facilitates the microglial response to amyloid plaque pathology, J Exp Med 215(4) (2018) 1047–1058.

38. T.K. Ulland, W.M. Song, S.C. Huang, J.D. Ulrich, A. Sergushichev, W.L. Beatty, A.A. Loboda, Y. Zhou, N.J. Cairns, A. Kambal, E. Loginicheva, S. Gilfillan, M. Cella, H.W. Virgin, E.R. Unanue, Y. Wang, M.N. Artyomov, D.M. Holtzman, M. Colonna, TREM2 Maintains Microglial Metabolic Fitness in Alzheimer’s Disease, Cell 170(4) (2017) 649–663 e13.

39. J.D. Ulrich, M.B. Finn, Y. Wang, A. Shen, T.E. Mahan, H. Jiang, F.R. Stewart, L. Piccio, M. Colonna, D.M. Holtzman, Altered microglial response to Abeta plaques in APPPS1-21 mice heterozygous for TREM2, Mol Neurodegener 9 (2014) 20.

40. S. Krasemann, C. Madore, R. Cialic, C. Baufeld, N. Calcagno, R. El Fatimy, L. Beckers, E. O’Loughlin, Y. Xu, Z. Fanek, D.J. Greco, S.T. Smith, G. Tweet, Z. Humulock, T. Zrzavy, P. Conde-Sanroman, M. Gacias, Z. Weng, H. Chen, E. Tjon, F. Mazaheri, K. Hartmann, A. Madi, J.D. Ulrich, M. Glatzel, A. Worthmann, J. Heeren, B. Budnik, C. Lemere, T. Ikezu, F.L. Heppner, V. Litvak, D.M. Holtzman, H. Lassmann, H.L. Weiner, J. Ochando, C. Haass, O. Butovsky, The TREM2-APOE Pathway Drives the Transcriptional Phenotype of Dysfunctional Microglia in Neurodegenerative Diseases, Immunity 47(3) (2017) 566–581 e9.

41. H. Keren-Shaul, A. Spinrad, A. Weiner, O. Matcovitch-Natan, R. Dvir-Szternfeld, T.K. Ulland, E. David, K. Baruch, D. Lara-Astaiso, B. Toth, S. Itzkovitz, M. Colonna, M. Schwartz, I. Amit, A Unique Microglia Type Associated with Restricting Development of Alzheimer’s Disease, Cell 169(7) (2017) 1276–1290 e17.

42. C. Sala Frigerio, L. Wolfs, N. Fattorelli, N. Thrupp, I. Voytyuk, I. Schmidt, R. Mancuso, W.T. Chen, M.E. Woodbury, G. Srivastava, T. Moller, E. Hudry, S. Das, T. Saido, E. Karran, B. Hyman, V.H. Perry, M. Fiers, B. De Strooper, The Major Risk Factors for Alzheimer’s Disease: Age, Sex, and Genes Modulate the Microglia Response to Abeta Plaques, Cell Rep 27(4) (2019) 1293–1306 e6.

43. N.M. Dräger, S.M. Sattler, C.T.-L. Huang, O.M. Teter, K. Leng, S.H. Hashemi, J. Hong, C.D. Clelland, L. Zhan, L. Kodama, A.B. Singleton, M.A. Nalls, J. Ichida, M.E. Ward, F. Faghri, L. Gan, M. Kampmann, A CRISPRi/a platform in iPSC-derived microglia uncovers regulators of disease states, bioRxiv (2021) 2021.06.16.448639.

44. C.M. Karch, A.M. Goate, Alzheimer’s Disease Risk Genes and Mechanisms of Disease Pathogenesis, Biol Psychiatry (2014).

45. R. Sims, S.J. van der Lee, A.C. Naj, C. Bellenguez, N. Badarinarayan, J. Jakobsdottir, B.W. Kunkle, A. Boland, R. Raybould, J.C. Bis, E.R. Martin, B. Grenier-Boley, S. Heilmann-Heimbach, V. Chouraki, A.B. Kuzma, K. Sleegers, M. Vronskaya, A. Ruiz, R.R. Graham, R. Olaso, P. Hoffmann, M.L. Grove, B.N. Vardarajan, M. Hiltunen, M.M. Nothen, C.C. White, K.L. Hamilton-Nelson, J. Epelbaum, W. Maier, S.H. Choi, G.W. Beecham, C. Dulary, S. Herms, A.V. Smith, C.C. Funk, C. Derbois, A.J. Forstner, S. Ahmad, H. Li, D. Bacq, D. Harold, C.L. Satizabal, O. Valladares, A. Squassina, R. Thomas, J.A. Brody, L. Qu, P. Sanchez-Juan, T. Morgan, F.J. Wolters, Y. Zhao, F.S. Garcia, N. Denning, M. Fornage, J. Malamon, M.C.D. Naranjo, E. Majounie, T.H. Mosley, B. Dombroski, D. Wallon, M.K. Lupton, J. Dupuis, P. Whitehead, L. Fratiglioni, C. Medway, X. Jian, S. Mukherjee, L. Keller, K. Brown, H. Lin, L.B. Cantwell, F. Panza, B. McGuinness, S. Moreno-Grau, J.D. Burgess, V. Solfrizzi, P. Proitsi, H.H. Adams, M. Allen, D. Seripa, P. Pastor, L.A. Cupples, N.D. Price, D. Hannequin, A. Frank-Garcia, D. Levy, P. Chakrabarty, P. Caffarra, I. Giegling, A.S. Beiser, V. Giedraitis, H. Hampel, M.E. Garcia, X. Wang, L. Lannfelt, P. Mecocci, G. Eiriksdottir, P.K. Crane, F. Pasquier, V. Boccardi, I. Henandez, R.C. Barber, M. Scherer, L. Tarraga, P.M. Adams, M. Leber, Y. Chen, M.S. Albert, S. Riedel-Heller, V. Emilsson, D. Beekly, A. Braae, R. Schmidt, D. Blacker, C. Masullo, H. Schmidt, R.S. Doody, G. Spalletta, W.T. Longstreth, Jr., T.J. Fairchild, P. Bossu, O.L. Lopez, M.P. Frosch, E. Sacchinelli, B. Ghetti, Q. Yang, R.M. Huebinger, F. Jessen, S. Li, M.I. Kamboh, J. Morris, O. Sotolongo-Grau, M.J. Katz, C. Corcoran, M. Dunstan, A. Braddel, C. Thomas, A. Meggy, R. Marshall, A. Gerrish, J. Chapman, M. Aguilar, S. Taylor, M. Hill, M.D. Fairen, A. Hodges, B. Vellas, H. Soininen, I. Kloszewska, M. Daniilidou, J. Uphill, Y. Patel, J.T. Hughes, J. Lord, J. Turton, A.M. Hartmann, R. Cecchetti, C. Fenoglio, M. Serpente, M. Arcaro, C. Caltagirone, M.D. Orfei, A. Ciaramella, S. Pichler, M. Mayhaus, W. Gu, A. Lleo, J. Fortea, R. Blesa, I.S. Barber, K. Brookes, C. Cupidi, R.G. Maletta, D. Carrell, S. Sorbi, S. Moebus, M. Urbano, A. Pilotto, J. Kornhuber, P. Bosco, S. Todd, D. Craig, J. Johnston, M. Gill, B. Lawlor, A. Lynch, N.C. Fox, J. Hardy, A. Consortium, R.L. Albin, L.G. Apostolova, S.E. Arnold, S. Asthana, C.S. Atwood, C.T. Baldwin, L.L. Barnes, S. Barral, T.G. Beach, J.T. Becker, E.H. Bigio, T.D. Bird, B.F. Boeve, J.D. Bowen, A. Boxer, J.R. Burke, J.M. Burns, J.D. Buxbaum, N.J. Cairns, C. Cao, C.S. Carlson, C.M. Carlsson, R.M. Carney, M.M. Carrasquillo, S.L. Carroll, C.C. Diaz, H.C. Chui, D.G. Clark, D.H. Cribbs, E.A. Crocco, C. DeCarli, M. Dick, R. Duara, D.A. Evans, K.M. Faber, K.B. Fallon, D.W. Fardo, M.R. Farlow, S. Ferris, T.M. Foroud, D.R. Galasko, M. Gearing, D.H. Geschwind, J.R. Gilbert, N.R. Graff-Radford, R.C. Green, J.H. Growdon, R.L. Hamilton, L.E. Harrell, L.S. Honig, M.J. Huentelman, C.M. Hulette, B.T. Hyman, G.P. Jarvik, E. Abner, L.W. Jin, G. Jun, A. Karydas, J.A. Kaye, R. Kim, N.W. Kowall, J.H. Kramer, F.M. LaFerla, J.J. Lah, J.B. Leverenz, A.I. Levey, G. Li, A.P. Lieberman, K.L. Lunetta, C.G. Lyketsos, D.C. Marson, F. Martiniuk, D.C. Mash, E. Masliah, W.C. McCormick, S.M. McCurry, A.N. McDavid, A.C. McKee, M. Mesulam, B.L. Miller, C.A. Miller, J.W. Miller, J.C. Morris, J.R. Murrell, A.J. Myers, S. O’Bryant, J.M. Olichney, V.S. Pankratz, J.E. Parisi, H.L. Paulson, W. Perry, E. Peskind, A. Pierce, W.W. Poon, H. Potter, J.F. Quinn, A. Raj, M. Raskind, B. Reisberg, C. Reitz, J.M. Ringman, E.D. Roberson, E. Rogaeva, H.J. Rosen, R.N. Rosenberg, M.A. Sager, A.J. Saykin, J.A. Schneider, L.S. Schneider, W.W. Seeley, A.G. Smith, J.A. Sonnen, S. Spina, R.A. Stern, R.H. Swerdlow, R.E. Tanzi, T.A. Thornton-Wells, J.Q. Trojanowski, J.C. Troncoso, V.M. Van Deerlin, L.J. Van Eldik, H.V. Vinters, J.P. Vonsattel, S. Weintraub, K.A. Welsh-Bohmer, K.C. Wilhelmsen, J. Williamson, T.S. Wingo, R.L. Woltjer, C.B. Wright, C.E. Yu, L. Yu, F. Garzia, F. Golamaully, G. Septier, S. Engelborghs, R. Vandenberghe, P.P. De Deyn, C.M. Fernadez, Y.A. Benito, H. Thonberg, C. Forsell, L. Lilius, A. Kinhult-Stahlbom, L. Kilander, R. Brundin, L. Concari, S. Helisalmi, A.M. Koivisto, A. Haapasalo, V. Dermecourt, N. Fievet, O. Hanon, C. Dufouil, A. Brice, K. Ritchie, B. Dubois, J.J. Himali, C.D. Keene, J. Tschanz, A.L. Fitzpatrick, W.A. Kukull, M. Norton, T. Aspelund, E.B. Larson, R. Munger, J.I. Rotter, R.B. Lipton, M.J. Bullido, A. Hofman, T.J. Montine, E. Coto, E. Boerwinkle, R.C. Petersen, V. Alvarez, F. Rivadeneira, E.M. Reiman, M. Gallo, C.J. O’Donnell, J.S. Reisch, A.C. Bruni, D.R. Royall, M. Dichgans, M. Sano, D. Galimberti, P. St George-Hyslop, E. Scarpini, D.W. Tsuang, M. Mancuso, U. Bonuccelli, A.R. Winslow, A. Daniele, C.K. Wu, C.A.E. Gerad/Perades, O. Peters, B. Nacmias, M. Riemenschneider, R. Heun, C. Brayne, D.C. Rubinsztein, J. Bras, R. Guerreiro, A. Al-Chalabi, C.E. Shaw, J. Collinge, D. Mann, M. Tsolaki, J. Clarimon, R. Sussams, S. Lovestone, M.C. O’Donovan, M.J. Owen, T.W. Behrens, S. Mead, A.M. Goate, A.G. Uitterlinden, C. Holmes, C. Cruchaga, M. Ingelsson, D.A. Bennett, J. Powell, T.E. Golde, C. Graff, P.L. De Jager, K. Morgan, N. Ertekin-Taner, O. Combarros, B.M. Psaty, P. Passmore, S.G. Younkin, C. Berr, V. Gudnason, D. Rujescu, D.W. Dickson, J.F. Dartigues, A.L. DeStefano, S. Ortega-Cubero, H. Hakonarson, D. Campion, M. Boada, J.K. Kauwe, L.A. Farrer, C. Van Broeckhoven, M.A. Ikram, L. Jones, J.L. Haines, C. Tzourio, L.J. Launer, V. Escott-Price, R. Mayeux, J.F. Deleuze, N. Amin, P.A. Holmans, M.A. Pericak-Vance, P. Amouyel, C.M. van Duijn, A. Ramirez, L.S. Wang, J.C. Lambert, S. Seshadri, J. Williams, G.D. Schellenberg, Rare coding variants in PLCG2, ABI3, and TREM2 implicate microglial-mediated innate immunity in Alzheimer’s disease, Nat Genet 49(9) (2017) 1373-1384.

46. S. Hsu, A.A. Pimenova, K. Hayes, J.A. Villa, M.J. Rosene, M. Jere, A.M. Goate, C.M. Karch, Systematic validation of variants of unknown significance in APP, PSEN1 and PSEN2, Neurobiol Dis 139 (2020) 104817.

47. S. Hsu, B.A. Gordon, R. Hornbeck, J.B. Norton, D. Levitch, A. Louden, E. Ziegemeier, R. Laforce, Jr., J. Chhatwal, G.S. Day, E. McDade, J.C. Morris, A.M. Fagan, T.L.S. Benzinger, A.M. Goate, C. Cruchaga, R.J. Bateman, N. Dominantly Inherited Alzheimer, C.M. Karch, Discovery and validation of autosomal dominant Alzheimer’s disease mutations, Alzheimers Res Ther 10(1) (2018) 67.

48. A.S. Mukadam, S.Y. Breusegem, M.N.J. Seaman, Analysis of novel endosome-to-Golgi retrieval genes reveals a role for PLD3 in regulating endosomal protein sorting and amyloid precursor protein processing, Cell Mol Life Sci 75(14) (2018) 2613–2625.

49. A.C. Gonzalez, M. Schweizer, S. Jagdmann, C. Bernreuther, T. Reinheckel, P. Saftig, M. Damme, Unconventional Trafficking of Mammalian Phospholipase D3 to Lysosomes, Cell Rep 22(4) (2018) 1040–1053.

50. L.C. Walker, Abeta Plaques, Free Neuropathol 1 (2020).

51. A.R. Ladiwala, J. Litt, R.S. Kane, D.S. Aucoin, S.O. Smith, S. Ranjan, J. Davis, W.E. Van Nostrand, P.M. Tessier, Conformational differences between two amyloid beta oligomers of similar size and dissimilar toxicity, J Biol Chem 287(29) (2012) 24765–73.

52. Y. Zhang, K. Chen, S.A. Sloan, M.L. Bennett, A.R. Scholze, S. O’Keeffe, H.P. Phatnani, P. Guarnieri, C. Caneda, N. Ruderisch, S. Deng, S.A. Liddelow, C. Zhang, R. Daneman, T. Maniatis, B.A. Barres, J.Q. Wu, An RNA-Sequencing Transcriptome and Splicing Database of Glia, Neurons, and Vascular Cells of the Cerebral Cortex, J Neurosci 34(36) (2014) 11929–47.

53. T.D. Troutman, E. Kofman, C.K. Glass, Exploiting dynamic enhancer landscapes to decode macrophage and microglia phenotypes in health and disease, Mol Cell 81(19) (2021) 3888–3903.

54. W.S. Liang, T. Dunckley, T.G. Beach, A. Grover, D. Mastroeni, D.G. Walker, R.J. Caselli, W.A. Kukull, D. McKeel, J.C. Morris, C. Hulette, D. Schmechel, G.E. Alexander, E.M. Reiman, J. Rogers, D.A. Stephan, Gene expression profiles in anatomically and functionally distinct regions of the normal aged human brain, Physiol Genomics 28(3) (2007) 311–22.

